# From emergence to endemicity: highly pathogenic H5 avian influenza viruses in Taiwan

**DOI:** 10.1101/2024.06.19.24309176

**Authors:** Yao-Tsun Li, Hui-Ying Ko, Joseph Hughes, Ming-Tsan Liu, Yi-Ling Lin, Katie Hampson, Kirstyn Brunker

## Abstract

A/goose/Guangdong/1/96-like (GsGd) highly pathogenic avian influenza (HPAI) H5 viruses cause severe outbreaks in poultry when introduced. Since emergence in 1996, control measures in most countries have suppressed local GsGd transmission following introductions, making persistent transmission in domestic birds rare. However, geographical expansion of clade 2.3.4.4 sublineages has raised concern about establishment of endemic circulation, while mechanistic drivers leading to endemicity remain unknown. We reconstructed the evolutionary history of GsGd sublineage, clade 2.3.4.4c, in Taiwan using a time-heterogeneous rate phylogeographic model. During Taiwan’s initial epidemic wave (January 2015 - August 2016), we inferred that localised outbreaks had multiple origins from rapid spread between counties/cities nationwide. Subsequently, outbreaks predominantly originated from a single county, Yunlin, where persistent transmission harbours the trunk viruses of the sublineage. Endemic hotspots determined by phylogeographic reconstruction largely predicted the locations of re-emerging outbreaks in Yunlin. The transition to endemicity involved a shift to chicken-dominant circulation, following the initial bidirectional spread between chicken and domestic waterfowl. Our results suggest that following their emergence in Taiwan, source-sink dynamics from a single county have maintained GsGd endemicity, pointing to where control efforts should be targeted to eliminate the disease.

## Introduction

Preventing emerging zoonotic viruses from establishing endemic circulation is essential to public health, considering the great health burden caused by endemic zoonoses globally^1^. An example of particular concern is the A/Goose/Guangdong/96-like (GsGd) H5 viruses, a lineage of highly pathogenic avian influenza (HPAI) first identified in southern China. From 2003-2006 HPAI GsGd viruses spread globally, infecting poultry in countries across Asia and Europe^2^. Most countries have subsequently eliminated HPAI GsGd viruses^3,4^, but, since 2010, a few countries, including Bangladesh, China, India, Indonesia, and Vietnam, have been classified as endemic, due to continued circulation in poultry and sporadic human cases^5^. Although there has been extensive characterisation of the genetic origins and the migration routes of HPAI GsGd viruses^6–8^, the mechanisms facilitating persistence and the transition to endemicity remain unknown.

Since emergence GsGd viruses have undergone diversification by accumulating mutations in their surface protein hemagglutinin (HA), resulting in multiple antigenically distinct sublineages, termed clades^2,3^. These genetic changes can alter host preferences, determining the virus’ reservoir, dissemination and persistence^9^. GsGd viruses of the clade 2.3.4.4, including clade 2.3.4.4a-2.3.4.4h, are distinguished from previous GsGd viruses by various neuraminidase (NA) subtypes (N2, N5, N6 and N8) besides N1, and have rapidly increased in the global population since 2014^10,11^. These "H5Nx" viruses have caused poultry and wild animal outbreaks in previously unaffected regions, such as the Americas^10,12^, and have also demonstrated unexpected transmission from Europe back to China^13^. Moreover, the viruses have led to mammal-to-mammal transmission in minks and cows^14,15^. The increasingly sustained circulation in areas previously thought to be sinks raises concerns about expanding establishment of endemic circulation^16^.

The clade 2.3.4.4c virus, one of the H5Nx sublineages, caused severe outbreaks in Korean poultry farms in early 2014^10,17^. Following migratory bird flyways, the virus spread to Japan, the USA and Taiwan by the end of that year^18–20^. In 2015, Taiwan experienced a devastating epidemic in domestic birds and culled over 5 million birds to curb localised outbreaks^21^. This marked the first significant transmission of HPAI GsGd viruses in the country^22,23^. While clade 2.3.4.4c viruses were eliminated in other countries before 2017^11,20^, the virus continued to circulate as H5N2 subtype in Taiwan until at least 2019^20,24^. With multiple genotypes generated by reassortment with local low pathogenic avian influenza (LPAI) viruses^20,23^, the HPAI clade 2.3.4.4c virus in Taiwan has been recognized as circulating endemically^21^. How this clade dispersed in Taiwan and the factors underlying its endemic establishment are not understood.

In this study, we quantified the persistence of GsGd outbreaks globally using virus HA genes, and identified the source of the endemic clade that established in Taiwan within this global context. We then curated the HA genes of the clade 2.3.4.4c virus in Taiwan, isolated from January 2015 to March 2019, representing approximately 20% of outbreaks that occurred within the country. Using this data set, we performed time-heterogeneous phylogeographic analyses to reconstruct the geographical and ecological dispersal dynamics of the clade as it transitioned from emergence to endemicity. Specifically, we elucidate the factors facilitating viral spread among counties/cities in Taiwan following the first epidemic wave in 2015.

## Results

### Global persistence of GsGd

To better understand local epidemics following GsGd introductions on a global scale, we identified descendent sublineages. We found about 400 sublineages from a uniformly downsampled dataset using the H5 genes of GsGd viruses up to September 2022 (Figure 1). These GsGd sublineages circulated across 78 countries, but their circulation was generally short-lived, with an average duration of 0.6 years based on the date of sample collection. Closer examination of sublineages that persisted for more than three years (based on a 95% quantile of 2.64), revealed 15 persistent sublineages in Southeast Asia, South Asia and the Middle East, including the clade 2.3.4.4c virus in Taiwan (Figure 1). Geographical reconstruction informed by outbreak reports supported this finding that most introductions had limited onward transmission (on average, 0.61 years) and identified the same countries with prolonged viral circulation (data not shown).

**Figure 1.**
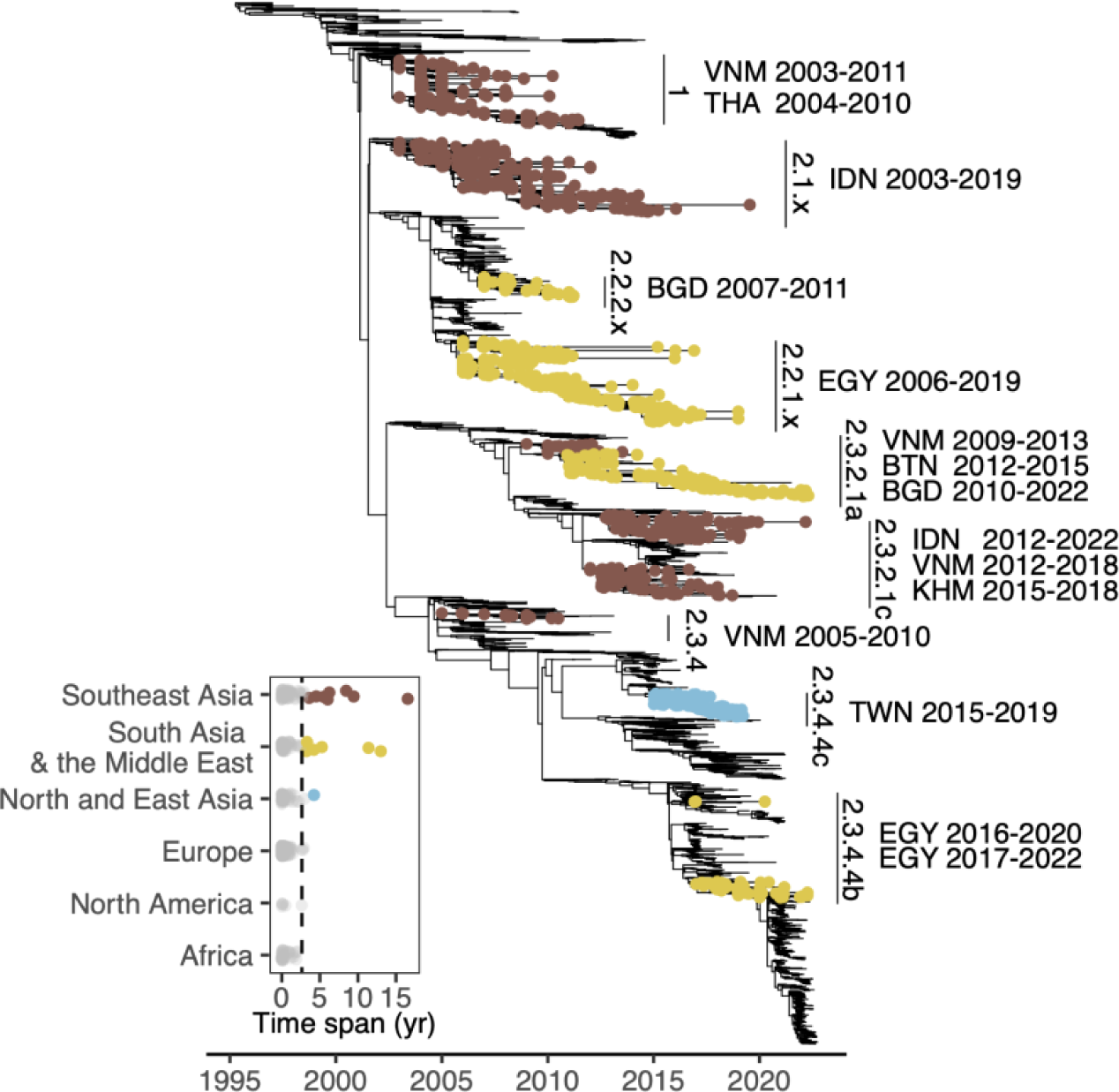
The duration of GsGd outbreak lineages. The time-scaled phylogenetic tree inferred with HA genes highlights fifteen lineages, which persistently circulated for at least three years. These are labelled with H5 nomenclature classification, country of isolation and duration. The inset shows the lineage duration classified by geographical area, with persistent sublineages highlighted as tips on the phylogeny. The dashed line denotes 95% quantile. VNM, Vietnam; THA, Thailand; IDN, Indonesia; BGD, Bangladesh; EGY, Egypt; BTN, Bhutan; KHM; Cambodia; TWN, Taiwan.

### The emergence and spread of clade 2.3.4.4c in Taiwan

In January 2015, over 600 outbreaks of clade 2.3.4.4c were reported in Taiwan across 10 counties/cities (Figure 2A). Subsequently, reported HPAI outbreaks in Taiwan declined and became sporadic in previously affected areas, with only three cases reported during July-August 2016, before transmission resumed in winter 2016-2017 (Figure 2B). To understand the spread of the clade 2.3.4.4c virus in Taiwan, we curated a genetic data set of viral HA sequences. The sequences are distributed over time from January 2015 to March 2019, and their sample sizes per county/city correlate well with the numbers of reported outbreaks (Figure 2D).

**Figure 2.**
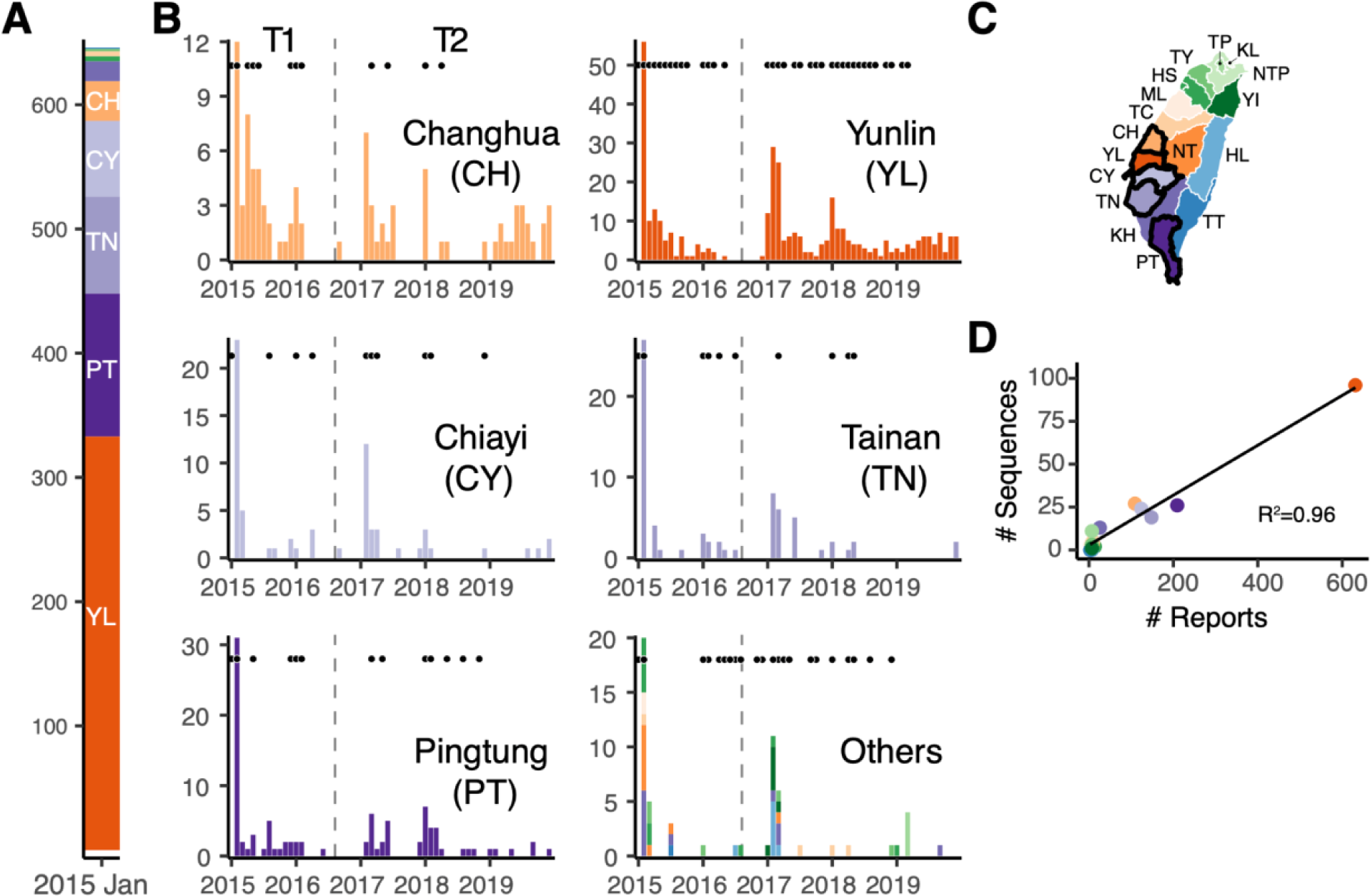
The emergence of HPAI GsGd clade 2.3.4.4c in Taiwan and subsequent circulation from 2015-2019. (A and B) The number of reports of infected poultry in farms or abandoned carcasses. The January 2015 numbers are pooled in panel A, while the monthly numbers of reports are presented by location in panel B. Black dots above the bars indicate the availability of genomic data in the corresponding months. The dashed vertical lines indicate the start of September 2016, the transition from the emergence (T1) to the endemic phase (T2). (C) The map highlights the five counties/cities with the most reports. (D) The correlation between reported outbreaks and available sequences for each of the 14 counties/cities. The points are coloured using the same colour scheme as the map and the other panels. NTP, New Taipei; TP, Taipei; KL, Keelung; TY, Taoyuan; HS, Hsinchu; YI, Yilan; ML, Miaoli; TC, Taichung; CH, Changhua; NT, Nantou; YL, Yunlin; CY, Chiayi; TN, Tainan; KH, Kaohsiung; PT, Pingtung; HL, Hualien; TT, Taitung.

To understand the dispersal dynamics of the 2.3.4.4c clade in Taiwan, we reconstructed the viral spread using continuous phylogeographical approaches which inferred the spatial locations of nodes. The Bayesian MCMC sampling process used priors informed by surveillance data to specify the locations of viruses (see Methods). The results, stratified by year, indicate that in 2015 there was intensive virus dispersal within and between western counties/cities (CH, YL, CY and TN), as well as more limited dispersal in southern counties/cities (KH and PT)(Figure 3). Long-distance dispersal events mostly originated from the main cluster in the west, and the northward dispersal events did not appear to seed sustained outbreaks. Since 2016, the number of dispersal events has significantly decreased and since 2017 Yunlin County (YL) has become the primary source of both short- and long-distance spread (Figure 3). The pattern is further illustrated by discretizing the inferred locations, with diffusion within YL dominating in all directions (Supplementary Figure 1).

**Figure 3.**
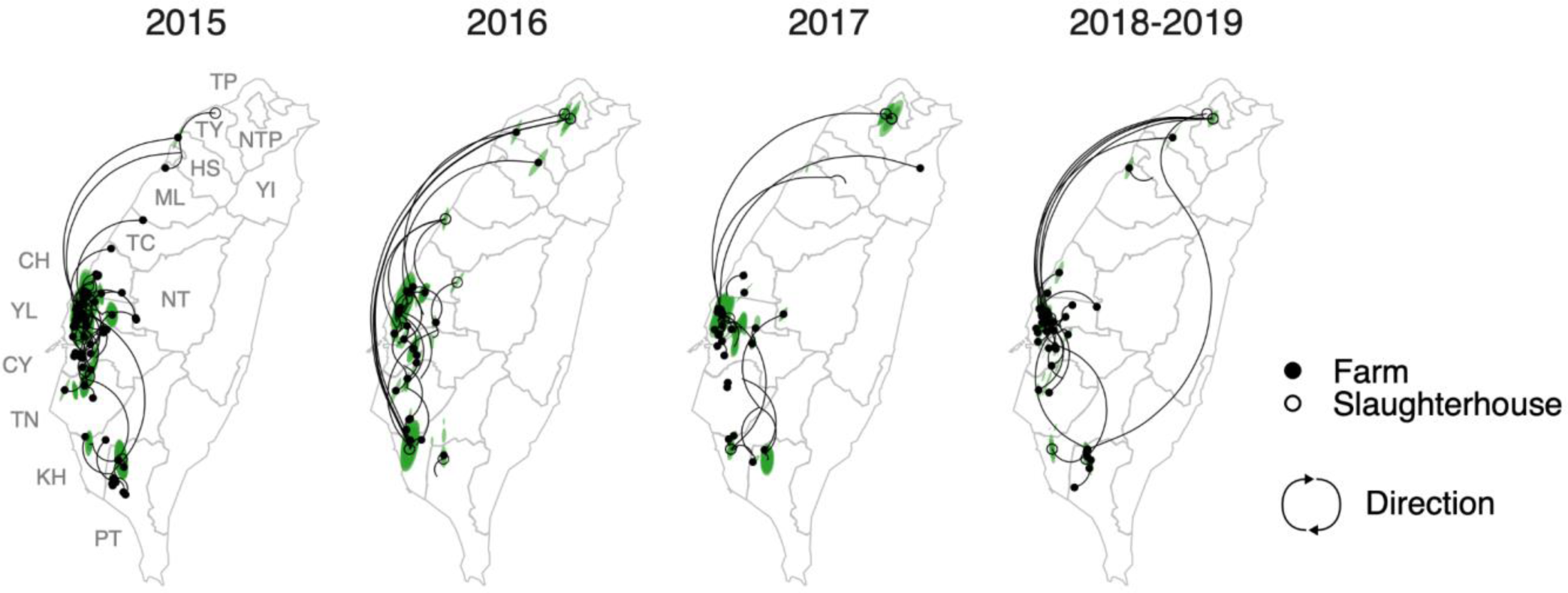
Reconstruction of the dispersal of clade 2.3.4.4c in Taiwan, 2015-2019. The points on the maps indicate the location of the virus samples, while the coloured areas represent the 80% HPD (highest posterior density) of the nodes inferred by the continuous phylogeography. Curves represent the branches of the maximum clade credibility (MCC) tree and are categorised into four periods based on the timing of parental nodes. Dispersal direction (clockwise) of the viral lineage is indicated by the curvature.

Use of explicit phylogeographic methods can reduce sampling biases^25,26^. Here, the locations of samples from slaughterhouses or rendering factories introduced inaccuracies into our inference. This is because these locations were unlikely to be the origins of viral spread, particularly those from the northern cities (e.g. TP and NTP) where agricultural activity is minimal (Supplementary Figure 2). To reconstruct the dispersal history of the virus, we therefore used discrete diffusion methods to reassign the locations of slaughterhouse samples. Specifically, we employed priors informed by the surveillance data, which represented possible sources of animals transported to slaughterhouses or rendering factories (Supplementary Figure 3B). Additionally, we used a heterogeneous rate model to identify changes in dispersal patterns between two epidemiological phases defined from the surveillance data (Figure 2): from January 2015 to August 2016 (T1) and from September 2016 onwards (T2).

The statistically supported routes that we inferred indicate a shift in dispersal from outbreaks having multiple sources to predominantly originating from a single source (Figure 4 and Supplementary Figure 4A), and are generally consistent with inference from the continuous phylogeographic analyses. During the emergence phase (T1), substantial transition events were detected between two western counties (YL and CH), in addition to dispersal from YL to nearby counties, with less frequent transitions identified from CH and PT to neighbouring counties/cities. In contrast, during the transition to endemicity (T2), all supported routes originated from YL, except for a less frequent path from PT. To account for local transmission, we quantified the maximum time interval assembled by branches with the same inferred state (Figure 4 and Supplementary Figure 5A). YL appears to be the only site where viruses persisted for more than 75% of both time periods (T1 and T2), suggesting its potential as a source of virus spread. Furthermore, when considering the viruses leading to the most recent isolates, CH harboured the “trunk” of the clade in 2015, but since 2016, YL has consistently been the major site harbouring viruses leading to the most distant progeny (Supplementary Figure 6A). These results suggest that YL has become the most prominent origin for nationwide dispersal following the initial epidemic wave. In contrast, circulation in other locations, particularly CH, has been largely suppressed, preventing spread to other counties/cities.

**Figure 4.**
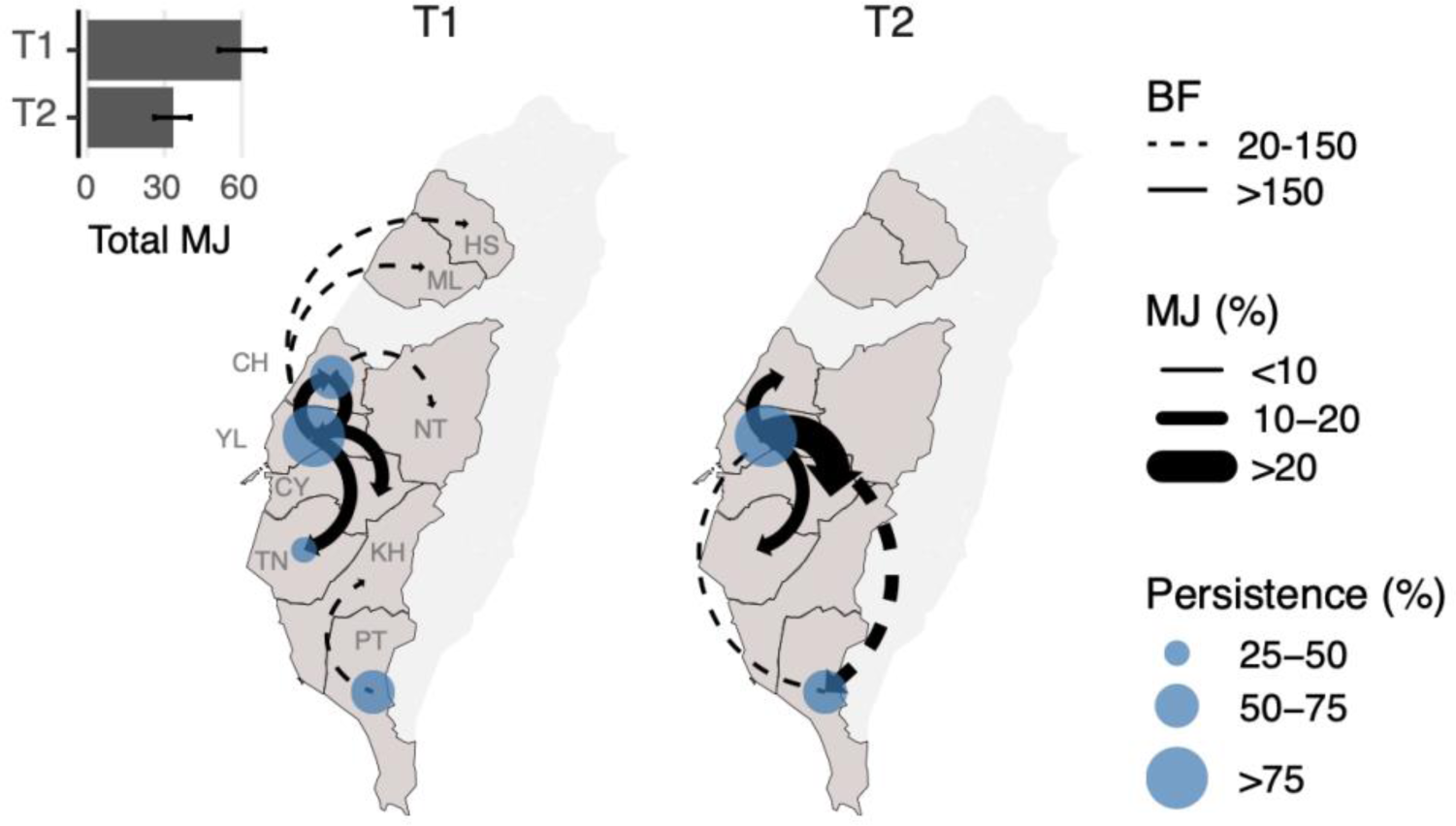
Schematic illustration of the main diffusion routes of the Taiwan clade 2.3.4.4c virus during the first epidemic wave and subsequent transition to endemicity. The statistically supported directions between counties/cities (Bayes factor>20) inferred by the discrete phylogenetic analyses are indicated as curves with arrows. The different line types reflect the strength of support (BF). The thickness of the curve reflects the frequency of transition events, presented as the proportion of total Markov jumps in the time period (T1 or T2). Total Markov jumps are indicated on the upper left inset. The size of the blue circles reflects the degree of persistence, calculated by unifying phylogeny branches with both nodes inferred as the same geographical area divided by the T1/T2 time period. Only areas where viruses circulated for more than 25% of the period are labelled. T1, January 2015 to August 2016; T2, September 2016 to March 2019.

To investigate the impact of sampling, we created two subsampled data sets by downsampling genetic sequences from county YL, resulting in YL no longer being the most frequently sampled site in each dataset. The subsampled data showed a shift in the origins of viral diffusion to CH or TN during the emergence phase (T1), but the inferred routes during the endemic phase (T2) were almost identical to those obtained using the full dataset (Supplementary Figure 4B and 4C). Similarly, persistence quantified from subsampling shows that CH replaced YL as the location with the most persistent circulation during the initial epidemic wave (T1). However, the estimates during the endemic phase (T2) were largely unaffected by the targeted subsampling (Supplementary Figure 5B and 5C).

To understand how the surveillance data may have biassed our inference, we also performed the same phylogeographic approaches based on the original collection county/city, with unknown samples assigned an uninformative prior containing all collection locations. This inference, which is independent of outbreak records, reveals a more widespread distribution of dispersal origins during the initial epidemic wave (T1). In addition to PT, more southern locations (i.e. TN and KH) were also identified as dispersal origins (Supplementary Figure 4D). During the endemic phase (T2), the pattern is consistent with the results obtained from considering the outbreak records. Notable differences were found in routes involving locations where samples were mainly from slaughterhouses or rendering factories (i.e. NTP and TP, Supplementary Figure 4D). Estimates of persistence based on record-independent priors (Supplementary Figure 5D) and all trunk probability conditions tested (Supplementary Figure 6B-D) indicate similar results to the full data set. Taken together, these findings suggest that our observation of nationwide geographical spread is robust to both sampling and state assignment.

### Location-specific factors drive the risk of viral dispersal

To identify the risk factors that facilitated the spread of clade 2.3.4.4c in Taiwan, we implemented a Generalised Linear Model (GLM) model accommodated by the time-heterogeneous discrete phylogeographic method. This allowed for estimates of predictor effect size and inclusion probability by time period. The results show that sample size is positively and significantly correlated with diffusion during the initial epidemic wave (T1, Figure 5A, RandEffect-). The number of poultry farms has a prominent inclusion probability during the endemic phase (T2), but this was not statistically significant. To account for location-specific factors, the GLM model was adjusted by adding random effects for each site, in addition to the existing predictors. The new model reduced the effects of sample size, and no further predictors were identified (Figure 5A, RandEffect+). Interestingly, the result shows that dispersal routes to high incidence counties/cities (CH, YL, CY and TN), and the direction from YL, have significant positive effects (Figure 5B). To avoid overparameterization, we conducted GLM estimations with only one of the predictors that had higher inclusion probabilities and sample size. Consistent with the full model, the reduced models that include predictors for either poultry farm, road distance, or cropland show no significant effects for these factors (Supplementary Figure 7). Furthermore, the random effects that specify directions to YL, CY, or TN are significant in all three reduced models, while the effect of the direction from YL is significant in one reduced model. The results remained consistent when the full model was examined in a time-homogeneous manner (Supplementary Figure 8). These findings suggest that location-specific factors are more likely to explain the spatial spread of the virus in Taiwan than shared agricultural risk factors.

**Figure 5.**
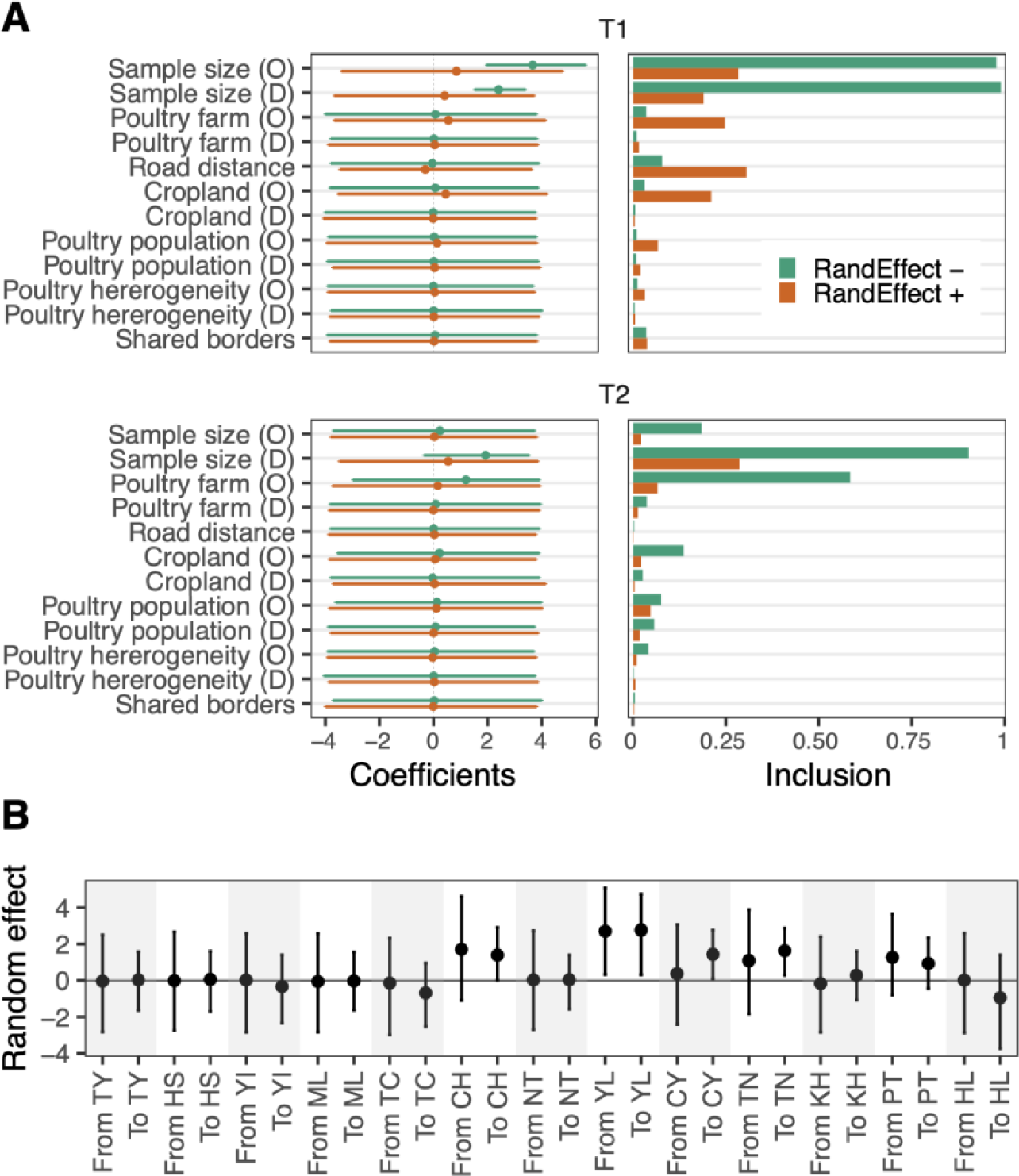
Potential predictors of dispersal of clade 2.3.4.4c virus between counties/cities in Taiwan. (A) The conditional effect sizes quantified by coefficients (left) and the inclusion probabilities of predictors (right) estimated by the time-heterogeneous phylogenetic generalised linear model (GLM). Results of the models with and without random effect variables are shown. The predictor names are denoted by O in parentheses for origin and D for destination. (B) Location-specific random effects in the GLM model. The effect sizes are in log space and are presented as mean with 95% HPD interval.

### Most YL outbreaks re-emerge from endemic hotspots

The phylogeographic results suggest a pivotal role of YL county in driving outbreaks across the country. However, the determinants that distinguish YL country from other locations remain unclear. To investigate this, we evaluated re-emerging HPAI outbreaks in the persistent foci during the endemic phase (T2). Based on the practice of establishing surveillance zones after case identification^21,27^, hotspots were defined by overlapping buffer areas within 1 km of node locations inferred by the continuous phylogeographic methods, or within the same distance of the node plus outbreak locations (Figure 6A). When mapping new outbreaks in YL from 2019 to 2022, over 50% of these occurred within the phylogenetically-defined hotspots. Moreover, over 70% of the outbreaks were found in the hotspots defined by the phylogenies supplemented with contemporary outbreak sites (Figure 6B). The areas defined by both methods account for less than 20% of the area of YL (Supplementary Figure 9). When the buffer radius was set to 0.5 km, over 30% and 60% of the new outbreaks were still identified within the hotspots defined by the two methods, accounting for less than 10% of the county’s area (Supplementary Figure 9). When the approach is applied to four other high incidence areas in Taiwan (Figure 2B), YL shows significantly higher percentages of recurrence in the hotspots defined by the two buffer distances (Figure 6C). These findings suggest that the inability to interrupt transmission within the county has led to the development of an endemic focus of persistent viral circulation in YL.

**Figure 6.**
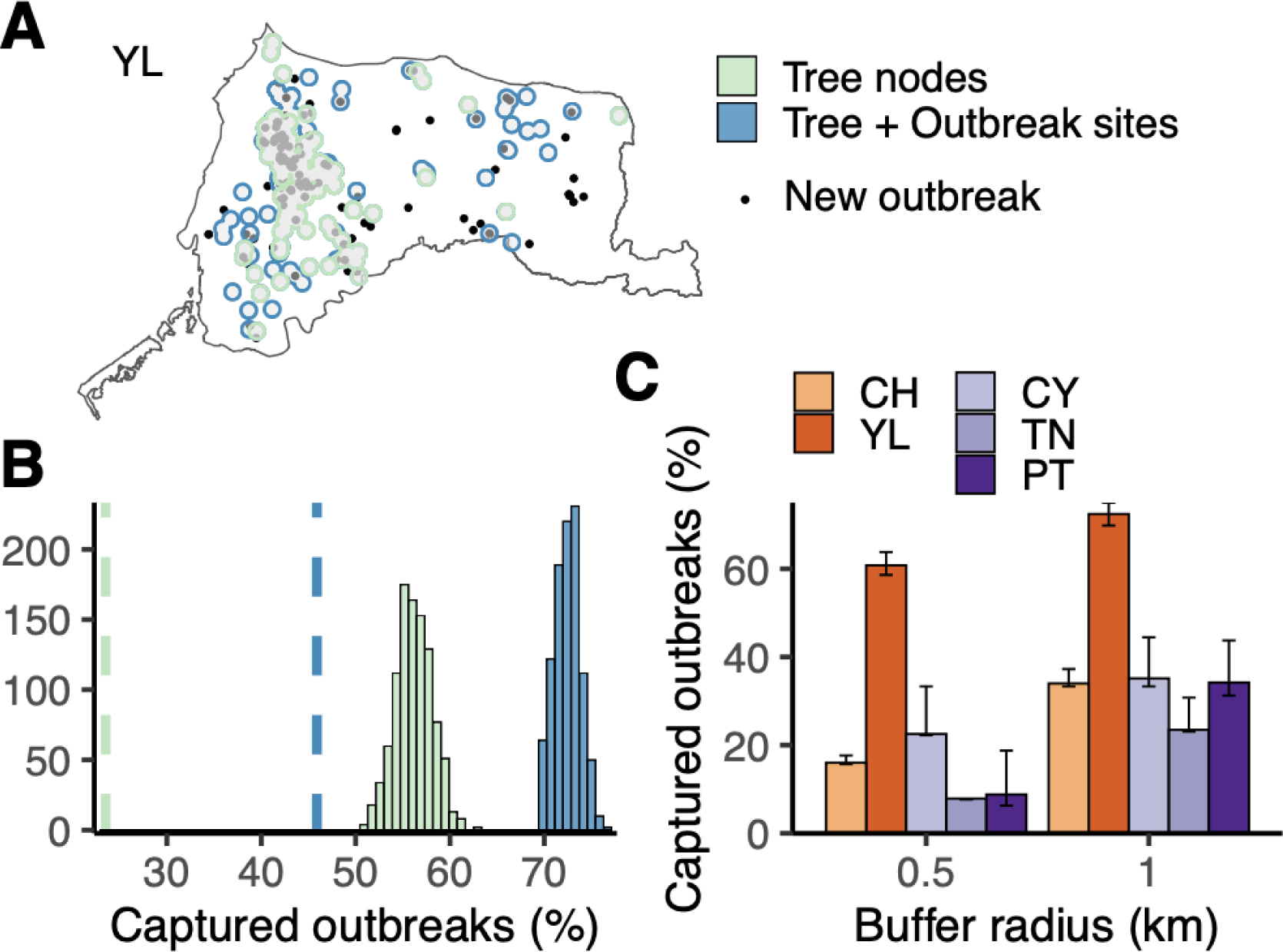
Evaluating the spatial distribution of re-emergent H5 HPAI outbreaks in Yunlin county (YL). (A) An example from a randomly selected posterior tree. The black points indicate reported outbreak sites that occurred between 2019 and 2022, after the latest available genetic data. The green/blue areas created by merged buffers indicate the inferred hotspots. (B) Distributions of captured new outbreaks by inferred epidemic hotspots based on 1 km-buffers with centres of nodes only (green) or nodes plus outbreak sites (blue). The vertical dashed lines indicate the mean of randomly generated circulating areas with identical buffer numbers as tree nodes (green) or tree nodes plus outbreak sites (blue). (C) Comparing the re-emergence of outbreaks within inferred hotspots in different locations. Proportions of captured outbreaks were calculated with buffers of two radius distances. Error bars indicate 95% credible intervals.

### Development of chicken-dominant circulation

The initial 2015 epidemic wave in Taiwan was characterised by widespread transmission in farms rearing domestic waterfowl^28^. This corresponded to a higher proportion of outbreaks and viral samples attributed to ducks or geese compared to chickens in 2015 (Figure 7B and Supplementary Figure 3A). To assess transmission between poultry species, we applied similar phylogeographic methods to the same genetic data, classified into Anseriformes (duck and goose) or Galliformes (chicken, turkey and quail) species. The inferred diffusion indicates that transmission between the two host groups was bidirectional during the initial epidemic wave (T1), with 80% of transmission attributed to spread from waterfowl, while both routes were statistically supported (Figure 7A). During the endemic phase (T2), transmission shifted to gallinaceous birds, while the transmission route from waterfowl was not supported (Figure 7A). This finding was further confirmed by an NA data set containing cognate N2 genes of Taiwan clade 2.3.4.4c viruses (Supplementary Figure 10). The trunk analyses show that the terrestrial poultry has maintained the gene source of the lineage since its introduction (Figure 7C). These results suggest that terrestrial poultry, mainly chickens, served as reservoir for the clade 2.3.4.4c virus during the endemic phase (T2), following the first epidemic wave during which inter-host group transmission dominated and more likely originated from domestic waterfowl.

**Figure 7.**
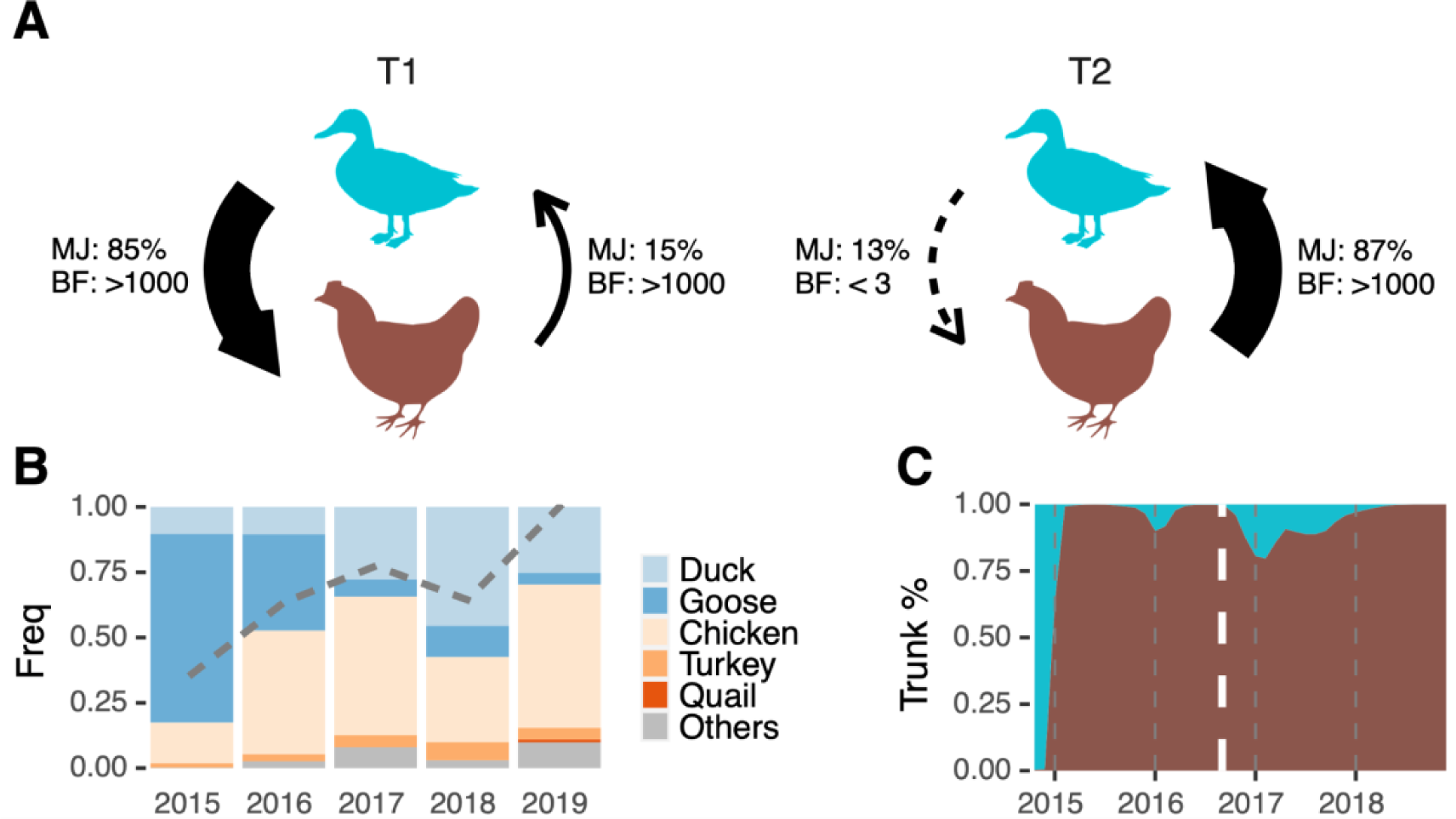
Transmission shifted from predominantly domestic waterfowl to chicken during the transition from epidemic to endemic circulation of 2.3.4.4c in Taiwan. (A) Shift in diffusion patterns between Anseriformes (duck and goose) and Galliformes (chicken, turkey and quail). Frequencies of transition events (Markov jump, MJ) and statistical support (Bayes factor, BF) were inferred using the discrete phylogenetic method. (B) The host distribution over time based on the surveillance data. The grey dashed line indicates the proportions of Galliformes in the genetic data. (C) Inferred host categories, i.e. Anseriformes or Galliformes species, of the trunk of clade 2.3.4.4c. The white dashed line divides T1 and T2, with areas coloured using the same scheme as panel (A).

By detecting signals of positive selection, we assessed the evidence for host adaptation. The dN/dS (w) estimate of HA (H5) genes in Taiwan clade 2.3.4.4c viruses is similar to that of HA genes in a North American H5N2 lineage introduced to the Taiwanese chicken population as early as 2003^20,29^, while the estimate of NA (N2) genes in the clade 2.3.4.4c viruses is higher than that of the North American lineage (Supplementary Table 1). In addition, there are less than five positively selected sites detected in both HA and NA of clade 2.3.4.4c in Taiwan, which is comparable to the results found in the North American H5N2 lineage. Note that all of the substitutions occurring at these sites were not fixed in the samples collected in 2019 (Supplementary Table 1). These results do not support selection pressure from host adaptation during the transition to endemicity and establishment of chicken-dominant circulation.

## Discussion

The circulation of clade 2.3.4.4c in Taiwan presents an opportunity to study how the GsGd lineage established endemic transmission following its emergence. By comprehensively characterizing the dynamics of clade 2.3.4.4c in Taiwan, this study focuses on viral dispersal and mechanisms driving endemic transmission after the initial epidemic wave. While the introduction and the genetic origins of the virus have been reported previously^20,23^, the dispersal dynamics and mechanisms driving persistence were previously unknown. Our time-heterogeneous phylogeographic analyses show that nearly all viral diffusion after mid-2016 originated from a single county, in contrast to the interconnected and multi-origin diffusion between counties/cities during emergence in 2015 (Figure 3 and 4). These endemic hotspots in Taiwan, where the disease was persistent, continued to seed outbreaks in other agricultural areas, while new outbreaks were mostly under control. Therefore, our geographical reconstruction provides insights into strategic control measures to eliminate GsGd viruses in Taiwan, emphasising the need to focus on the endemic hotspots. Note that the clade 2.3.4.4c virus was still being identified in 2023^24^, although viral genetic data isolated after 2019 was unavailable.

Previous studies using phylogenetic GLMs have identified the poultry trade^30^, geographic distance^31^ and the wild bird migration network^13^ as predictors of GsGd viral dispersal. These studies have investigated transmission patterns globally as well as single epidemic waves, but have not examined the risk of establishment of endemic transmission. Our GLMs did not identify any significant agricultural predictors for GsGd viruses in Taiwan. Instead, our analyses highlighted location-specific factors that were not explained by the tested predictors. These results indirectly support nationwide spread from a specific endemic county with other risk factors playing only a minor role. It is possible that overparameterization due to sample size could not be excluded, although different models were extensively explored (Supplementary Figures 7-8). Of all the predictors tested in the GLMs, the number of poultry farms had the highest inclusion probability (Figure 5 and Supplementary Figures 7-8), a finding corroborated by a spatial case-control study focusing on clustering of H5 viruses in Taiwan from 2015-2017^27^.

Lineage-specific host preferences can be targeted to prevent viral spread^32^. Control measures for HPAI have targeted domestic waterfowl due to their biological similarity to wild waterfowl, which are capable of long-distance dissemination^8,33^. In addition, infected waterfowl exhibit mild clinical manifestations compared with Galliformes species like chickens, increasing the risk of silent transmission and co-infection with multiple strains^2^. Our diffusion analyses indicate that GsGd transmission in Taiwan shifted from bidirectionality, with mixing between domestic waterfowl (Anseriformes species) and terrestrial species (Galliformes species), to a predominantly chicken-origin circulation (Figure 7). The greater transitions from waterfowl to terrestrial species during the first epidemic wave, aligned with devastating outbreaks in geese^21,28^, that did not lead to sustained descendant lineages (Figure 7C). Based on the selection analyses of the surface proteins, it is unlikely that the change in dispersal patterns between these epidemiological phases reflects virus adaptation from waterfowl to terrestrial birds (Supplementary Table 1). Instead, the results suggest that unknown factors in the chicken farming networks supported endemic circulation of GsGd in Taiwan, while viral lineages circulating in goose and duck populations were mostly eliminated. During the transition to endemicity, more than half of the Anseriformes samples were either isolated or inferred to be from Yunlin county (Supplementary Figure 3), indicating that the diffusion pattern in host ecology was not confounded by the spatial pattern described earlier.

Although these data have provided invaluable information on H5 HPAI circulation in Taiwan, it is unlikely that the surveillance data from the Taiwanese agricultural authorities identified all of the GsGd or clade 2.3.4.4c outbreaks. This limitation is implied by the results of the discrete diffusion analysis that relied solely on genetic sequences without outbreak records (Supplementary Figure 4D). The analysis exclusively identified statistically supported transmission routes originating from locations in southern Taiwan, particularly city KH, during the first epidemic phase. It was also noted that some viral samples do not precisely correspond to inferred locations (Figure 2B and Methods). Our reconstruction schemes demonstrate that state assignment strategies have little effect on all tested characteristics during the endemic phase, T2 (Supplementary Figure 4-6). There may also be an issue with the lack of explicit viral classification for each entry in the surveillance data, leading to inaccurate priors for the locations of the 2.3.4.4c virus. However, there is no evidence of continued circulation of other GsGd lineages, including clade 2.3.4.4b, in Taiwan during 2015-2019, according to available sequences (see Methods) and outbreak reporting^24^.

Our findings indicate that in Taiwan, HPAI control should prioritise the source region, specifically by strengthening measures to interrupt transmission and improve biosecurity in poultry farms in Yunlin county. Meanwhile, epidemics in other regions could be readily suppressed. On a global scale, attention should be paid to countries newly affected by GsGd viruses after 2020^12,16^, as well as countries where viruses were previously seeded (Figure 1). Effective interventions can be implemented through continued local and global surveillance efforts to discern dispersal patterns supported by genomic data.

## Materials and Methods

### Sequence data preparation

H5 hemagglutinin (HA) sequences of avian influenza A viruses were downloaded from NCBI (Influenza Virus Resource, https://www.ncbi.nlm.nih.gov/genomes/FLU/)^34^ and GISAID (EpiFlu, https://www.gisaid.org)^35^ on 17 October, 2022. N2 neuraminidase (NA) sequences of part of the associated H5 viruses in Taiwan were also downloaded from the same databases. Short sequences (<1500 for HA and <1200 for NA) or sequences containing notable (n=100) ambiguous nucleotides were removed from the download. Filtered genetic data were then aligned using Nextalign v2.3.0^36^ or MAFFT v7.490^37^ and trimmed to coding regions. Multiple basic amino acids at the H5 cleavage site were also removed. Sequences of Taiwan GsGd clade 2.3.4.4c viruses analysed in this study have all been described previously^20,21,23,28,38,39^.

### Quantifying the persistence of GsGd lineages globally

The above GsGd H5 sequences were first subjected to location-focused downsampling. That is, for each country where genetic data were isolated, five sequences were randomly sampled on different dates within a monthly interval. When countries had samples with incomplete temporal information, up to 50 sequences were allowed to be randomly included per year. The downsampled data set was then used to reconstruct a time-scaled tree using Treetime v0.11.1^40^ and a maximum likelihood (ML) tree inferred by IQ-TREE v2.2.0^41^, justified by a strong temporal signal (R^2^=0.95). The *mugration* function in Treetime was performed to infer the country as ancestral states for each node. Post-introduction sublineages were defined by first identifying internal branches representing state transition and all the descending taxa of those branches. For each of these sublineages, nested transition branches were removed, resulting in a lineage each comprising taxa isolated in the same country and sharing a most common ancestor linked to viral movement. Considering the variation in sequencing effort across countries, ancestral reconstruction based on transition rates informed by frequencies of HPAI H5 outbreaks during 2000-2022^42^ was also performed.

### Phylogenetic analyses and data set design

A ML tree was inferred for the HA alignment using IQ-TREE. Major H5 viral lineages including clades 2.3.4.4b and c, were classified according to the GsGd H5N1 nomenclature proposed by the World Health Organization^3,11^. All available H5 viruses isolated in Taiwan were identified on the tree. A total of 292 HA sequences isolated in Taiwan were classified as clade 2.3.4.4c, accounting for nearly all the available GsGd isolates in the country (292/295). The main working dataset was generated by removing duplicated sequences and viruses isolated in apparently the same outbreak (n=252). To verify results in the discrete phylogeographic analyses, more evenly distributed datasets were prepared by downsampling the sequences from Yunlin county (YL). The downsampled datasets, which include no more than six samples per year in YL, contain comparable sample sizes as the other locations where disease was prevalent (sample size in county YL, 26; CH, 27; CY, 24; PT, 26). In addition, to verify patterns in host diffusion, the N2 genes corresponding to the curated H5 datasets were collected (n=183); N2 is the predominant subtype among clade 2.3.4.4c viruses in Taiwan (Supplementary Figure 3). Temporal signal in each data set was examined by TempEst v1.5.3^43^, taking an ML tree inferred by IQ-TREE and times of sample collection as input. All phylogenetic trees in this study were visualised with the ggtree package^44^.

### Epidemiological and spatial information of HPAI H5 viruses in Taiwan

The Taiwanese government has been actively conducting virological surveillance of HPAI H5 viruses in poultry farms and abandoned bird carcasses prior to 2015^28,39^. Information on the collection date, host species and subtypes of each identification report can be found on a publicly available website^24^. The sampling sites’ geographical coordinates for the reported events were obtained by parsing an interactive map embedded on the same website using the RSelenium package^45^. Metadata for the genetic data, including subtype, time, location at the county/city level and host, were acquired along with the sequences. Information on the sampling environment types, i.e., farms or slaughterhouses, were obtained from a recent publication by the Taiwanese government^21^.

### Continuous phylogeographic analyses

We adopted a recently developed method to account for uncertainty in the sampling location for evolutionary reconstructions in continuous space^46,47^. Specifically, for each sequence, a prior describing multiple polygons and their corresponding probabilities was defined in a Keyhole Markup Language (KML) file, and thereby incorporated into the Bayesian Markov chain Monte Carlo (MCMC) process performed by BEAST^48^. To construct the priors in this study, candidate locations for each genetic sequence were defined based on the sampling sites of the surveillance data. The candidate locations were gathered by matching the sequence’s metadata, i.e. host, subtype, and county/city, to the reported events, along with a dynamic time-interval starting with (1) the collection date; if no hit was found with compatible information and in the time-interval, (2) the collection date +-7 was used. After conducting two searches, the centroids of all subdivisions in the county/city (i.e. town or area) of the sample were assigned to each unmatched sequence. The probabilities of a prior were uniform for sequences that were compatible with events in the surveillance data. However, for sequences that we were unable to link to reported events, the probabilities were assigned proportionally to the incidence of H5 HPAI during 2015-2019 in subdivisions. Each site with geographical coordinates was expanded to a minimum area of latitude +-0.0003 and longitude +-0.0005 to fit the polygon format in the original approach^46^. A sample collected in a slaughterhouse or rendering factory was assigned to the location of one arbitrarily selected slaughterhouse for each county/city based on the registry data (www.baphiq.gov.tw, accessed 22 March 2023).

Continuous phylogeographic analyses were performed with the Cauchy Relaxed Random Walk (RRW) model implemented in BEAST v1.10.4^25,49^. A general time reversible (GTR) substitution model, a gamma-distributed rate and an uncorrelated lognormal relaxed molecular clock^50^ were employed in the Bayesian analyses, along with the Skygrid demographic model^51^. The MCMC was run for 550 million steps, with samples taken every 10,000 steps after removing 50 million steps as burn-in. Parallel runs were conducted to confirm the observed pattern. A starting tree was added to each run to facilitate the sampling of tree topologies. The results of continuous phylogeographic analyses were visualised with the SERAPHIM package^52^.

### Discrete phylogeographic analyses with an epoch model

To evaluate differences in spatial dispersal patterns throughout different epidemiological phases, from the introduction to the development of endemicity, we employed a time-heterogeneous transition rate model with the discrete phylogeographic method^53^. The method enables the Bayesian discrete state inference to accommodate multiple rate matrices, each of which specified by one time interval (epoch). Therefore, transition frequencies and statistically supported directions in different intervals can be simultaneously estimated on the same phylogeny. We defined two time intervals, T1 and T2, divided by the date 1 September, 2016, based on sample sizes of the sequence data and the epidemiological trajectory (Figure 2). Transition parameters were estimated using an asymmetric substitution model with Bayesian stochastic search variable selection (BSSVS)^54^.

To better approximate the locations of the original farms, uncertain discrete state assignment was applied to the samples collected in slaughterhouses or rendering factories^55^. Similar to the strategy we used for the analyses in continuous space, we informed the priors of the uncertain discrete locations with surveillance data. We gathered candidate locations at the county/city level by matching the sequence’s metadata to the reports within a 15-day interval (collection date +-7). If no matched events were found, we extended the interval to 90 days. The probabilities linked to the locations were proportional to the corresponding events in different counties and cities. The dispersal between hosts was also assessed by classifying sequences into the order of Anseriformes or Galliformes.

Additionally, we used a generalised linear model (GLM) incorporated into the discrete phylogenetic framework to investigate the potential predictors’ contribution to the transition between locations^56^. The predictors included (1) road distance, (2) shared border, (3) number of poultry farms, (4) poultry population, (5) poultry heterogeneity, (6) area of cropland and (7) number of viral sequences. Poultry farm, poultry heterogeneity and cropland area were added to the model as covariates for both origin and destination. Poultry farm, poultry population, poultry heterogeneity and sample size were treated as time-heterogeneous by averaging the annual values in the two intervals (2015-2016 and 2016-2019)(Supplementary Figure 2), while all predictors were log-transformed and standardised before inclusion in the matrices. The GLM model was adapted using previously described methods to assume temporally heterogeneous effect sizes and inclusion probabilities^13,57^. To account for unexplained variability, models that included time-homogeneous random effect variables specifying the effects as both origin and destination of each location were also implemented^58,59^. To validate the results, we performed reduced GLM models with no more than two predictors and a time-homogeneous GLM model including all the aforementioned predictors.

BEAST v1.10.5 (pre-released v0.1.2) was used to implement time-heterogeneous rate models with MCMC chains of 110 million steps^48^. Performance of the GLM estimations were improved by applying a set of empirical trees. All BEAST runs were facilitated by the BEAGLE library^60^. Convergence was examined using Tracer v1.7.2^61^, ensuring that effective sampling sizes (ESS) were greater than 200 for all continuous parameters.

Posterior analyses. The number of transitions between the two states was quantified by Markov jumps^62^. Bayes factors (BF) were calculated based on the indicators of transition rates resulting from BSSVS to determine statistically supported diffusion routes. Routes with BF>20 were considered supported and were classified into two categories: 20-150 and >150, which are typically referred to as ’strong’ and ’very strong’^63^. The duration of viral persistence was evaluated using maximised time intervals that were unified by branch lengths, where both nodes were inferred to be in the same location. Trunk probabilities over time were summarised by PACT v0.9.4 (https://github.com/trvrb/PACT). Trunks were defined as ancestral branches shared by sequences isolated in 2019. Persistence and trunk probability were calculated using 1000 MCMC trees sampled from the posterior phylogeny distributions.

### Probing the spatial distribution of re-emergent outbreaks

Buffers representing disease hotspots on the map were created by parsing geographical coordinates inferred at the nodes of the posterior trees in the continuous phylogeographic calibrations, including tips and internal nodes. A buffer was defined as the area within 1 km of the inferred point location. The buffers created by the phylogenetic inference were merged with or without the buffers created by the sampling sites of contemporary outbreak events reported in the surveillance data for each tree. Only nodes estimated no earlier than 1 September, 2016 (i.e. during the endemic phase, T2) and the outbreaks during the same period were taken as central points to create buffers. The nodes and the outbreaks were stratified by county/city before being transformed from points to polygons. Coverage ("captured") was determined by the sites of new outbreaks covered by the merged buffer areas. New outbreaks were defined as outbreaks occurring after the latest genetic sample, during 2019-2022. The captured outbreaks were summarised as proportions with 1000 posterior trees, whose node numbers were also used to generate randomly distributed null models. To evaluate the coverage performance, sensitivity analyses were conducted on buffer size considering multiple radius distances, including 0.5, 1 and 2 km. The spatial data manipulation was performed using the *sf* package^64^. The map data (shp files) were obtained from the government open data platform (data.gov.tw).

### Data sources of potential predictors

Road distances between counties and cities in Taiwan were measured with Google Maps. Data on agricultural statistics, including the number of poultry farms, poultry population and cropland area during 2015-2019 were obtained from the Agricultural Statistics Database of the Ministry of Agriculture (https://agrstat.moa.gov.tw/sdweb). Poultry heterogeneity was calculated as Simpson’s Index of Diversity^65^.

### Selection analyses

SLAC^66^ and MEME^67^ in the Datamonkey platform^68^ were used to detect pervasive and episodic positive selection sites, respectively, with a default statistical significance level of p<0.1. HyPhy v.2.3.14^69^ was used to calculate non-synonymous/synonymous rate ratio, dN/dS (ω) estimates. The results of the surface proteins in the clade 2.3.4.4c viruses were compared with those in an enzootic, non-GsGd H5N2 lineage previously described in Taiwan^20,29^.

### Data and code availability

The custom codes used in anaylses, and the XML files required for BEAST can be found at https://github.com/yaotli/endemic. The accession numbers of the Taiwan sequences analysed in our study are listed in Supplementary Table 2.

## Data Availability

The custom codes used in analyses, and the XML files required for BEAST can be found at https://github.com/yaotli/endemic. The accession numbers of the Taiwan sequences analysed in our study are listed in Supplementary Table 2.

## Acknowledgements

The work was supported by the UK Research and Innovation Global effort on COVID-19 (MR/V035444/1), Wellcome (207569/Z/17/Z, 224520/Z/21/Z), the UK Medical Research Council New Investigator Research Grant (MR/X002047/1) and a University of Glasgow Lord Kelvin/Adam Smith Fellowship to KB. We are grateful for the continued surveiilnace efforts of the Animal and Plant Health Inspection Agency, MOA, Taiwan. We acknowledge the authors, originating laboratories, and submitting entities responsible for providing the sequences obtained from GISAID.

## Author contributions

Conceptualisation: YTL, HYK. Formal analysis: YTL. Methodology: YTL, JH. Resources: HYK, YLL. Writing - original draft preparation: YTL. Writing - review & editing: YTL, HYK, JH, MTL, YLL, KH, KB. Supervision: JH, KH, KB. Funding: KH, KB.

---

**Supplementary figure 1.**
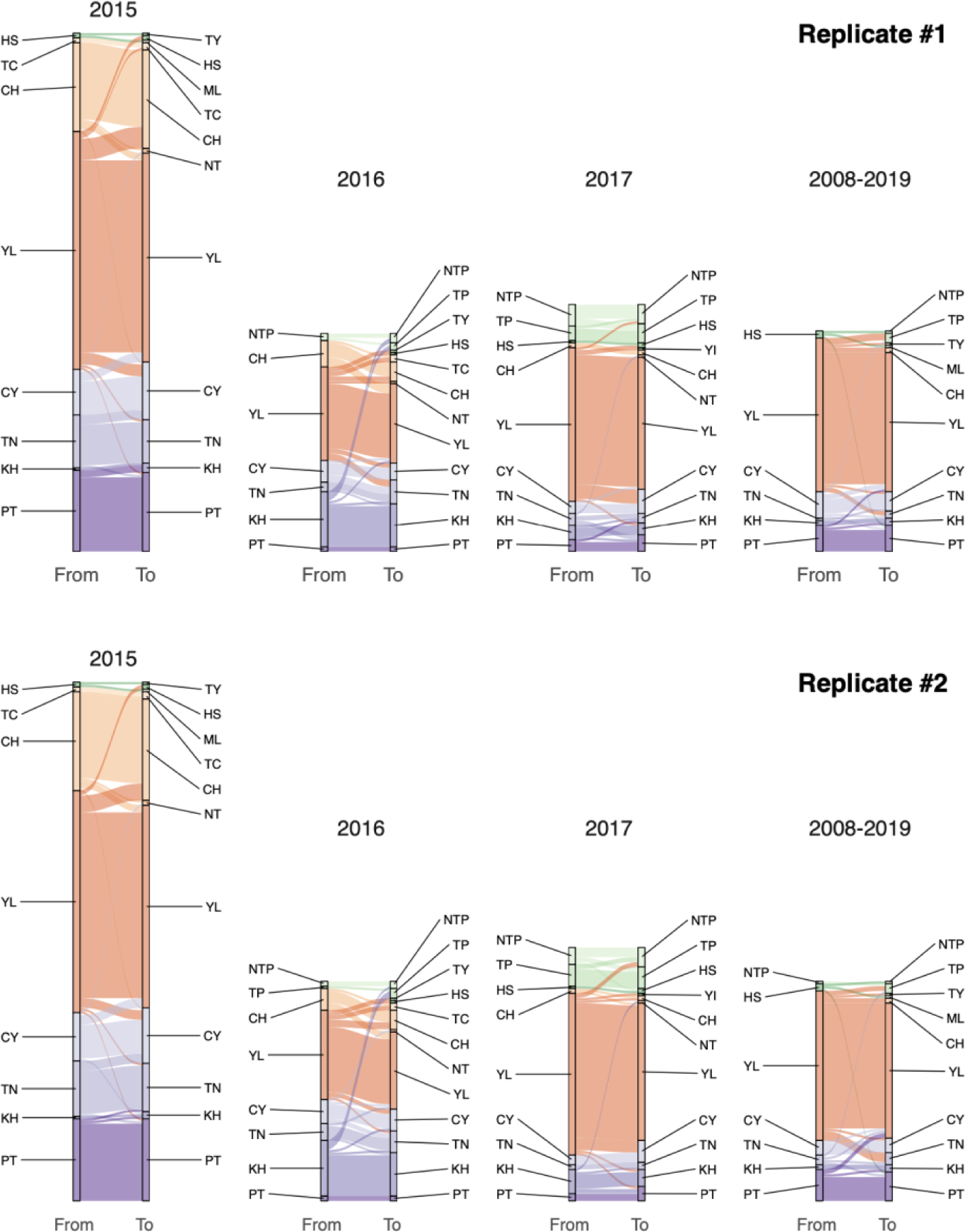
Visualisation of the dispersal pattern of the clade 2.3.4.4c virus in discrete geographical divisions. The connected lines represent branches inferred by the continuous phylogeographic approach (see Figure 3), with origin and destination locations determined as county or city-level divisions. The lines were coloured based on the origin. Replicated results from parallel runs are shown.

**Supplementary figure 2.**
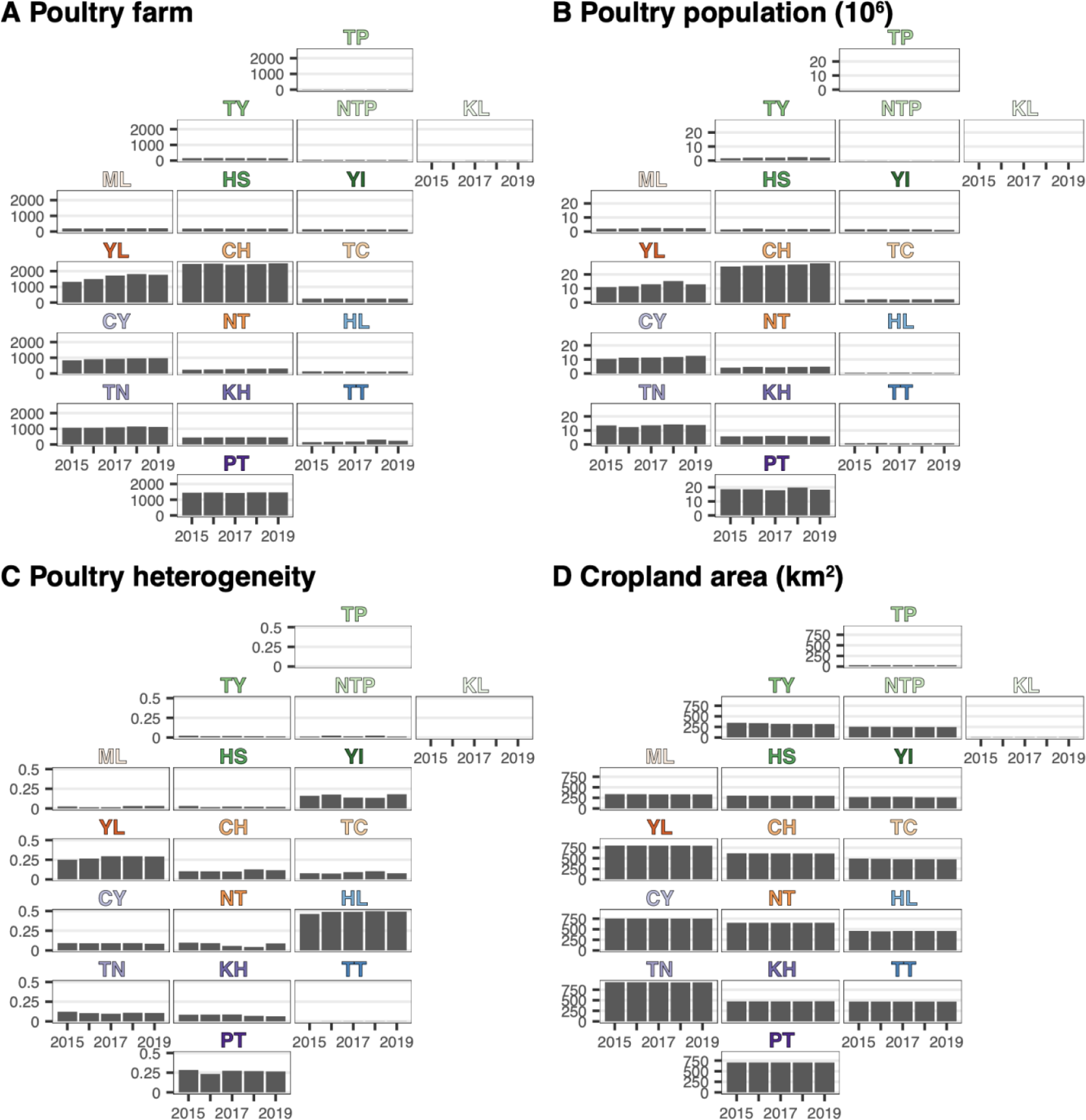
Agricultural characteristics of county or city-level administrative areas in Taiwan.

**Supplementary figure 3.**
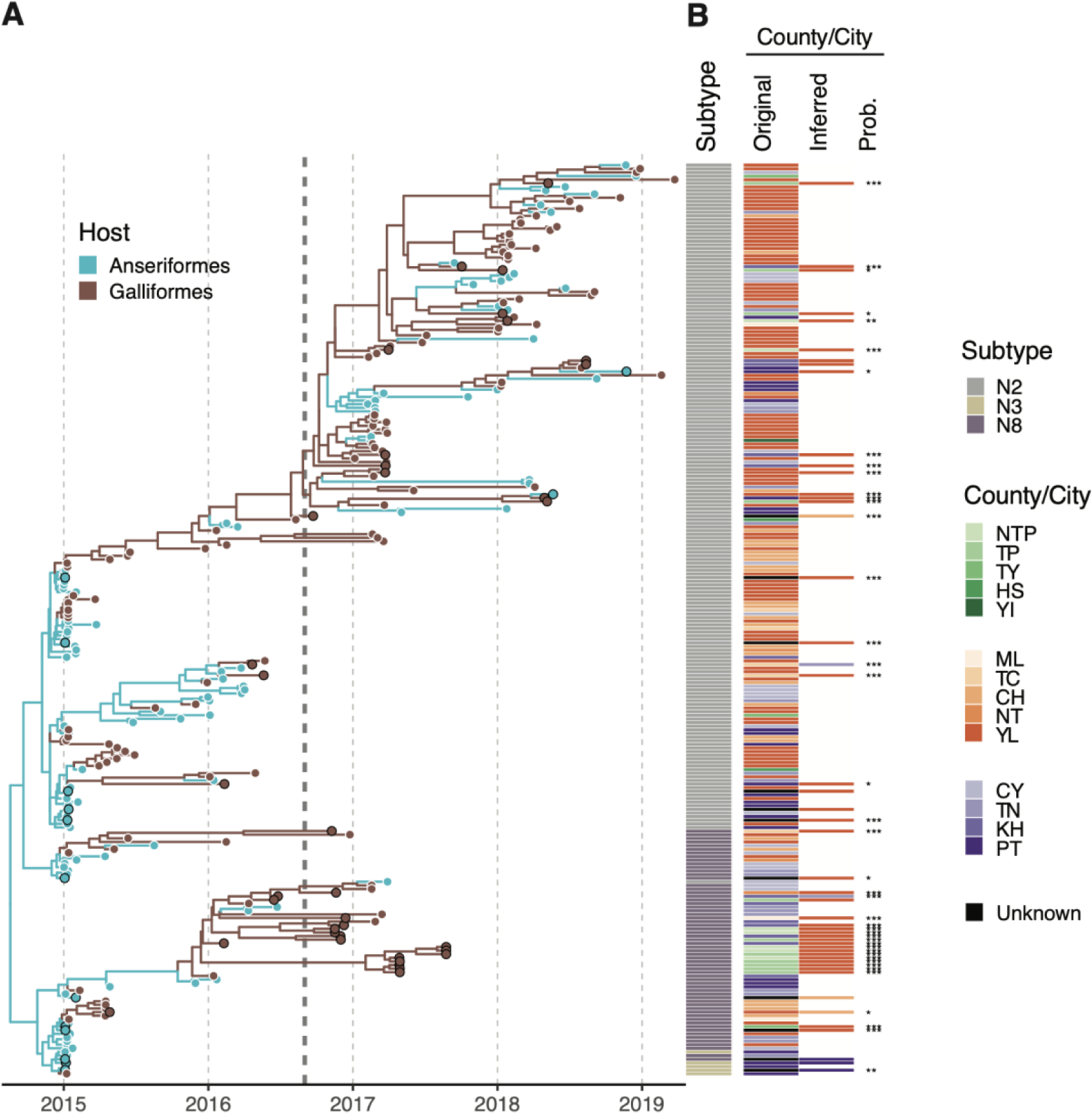
Summary of the host and geographical information associated with the genomic data used in this study. (A) The time-scaled phylogeny was reconstructed using the HA genes of the clade 2.3.4.4c virus in Taiwan. The tips on the phylogeny are colored according to the host genera, Galliformes (chicken, turkey and quail) or Anseriformes (duck and goose), logged in the sequence metadata. The branches are colored according to the states inferred on the summarized maximum clade credibility (MCC) tree. The tips with black borders indicate taxa that have uncertain state assignments in geographical analyses. The vertical dashed line denotes September 1st, 2016. (B) The adjacent heatmap shows the NA subtypes and the corresponding collection locations of the tree taxa. The discrete diffusion model was used to estimate the uncertain geographical locations and posterior probabilities. Only the results of samples collected in slaughterhouses/rendering factories and samples lacking geospatial information, which were implemented with state uncertainty, are shown in the ’Inferred’ column. The symbols *, ** and *** denote posterior probability>0.7, 0.8 and 0.9, respectively.

**Supplementary figure 4.**
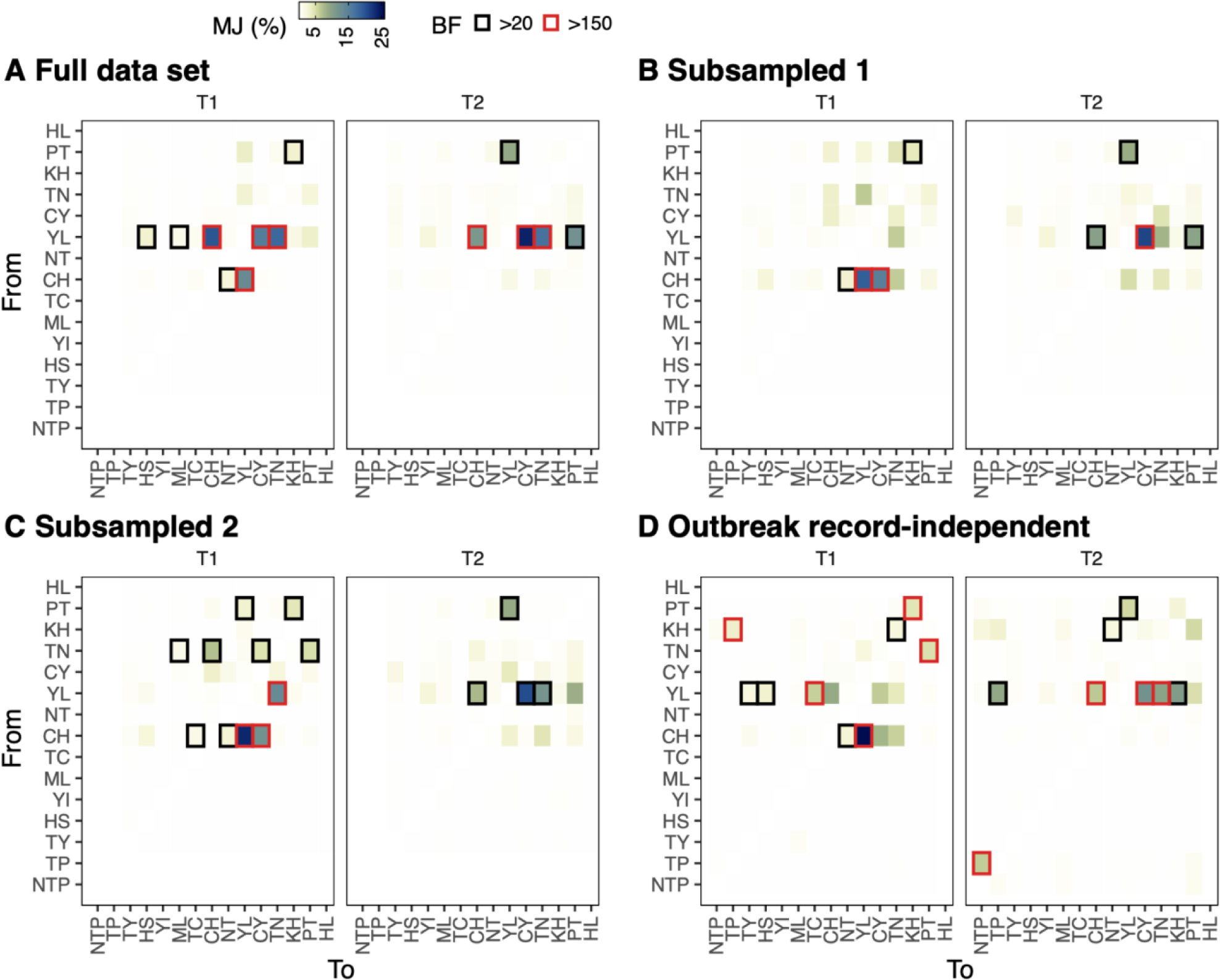
Estimated number of transition events between counties/cities in Taiwan inferred by the discrete phylogeographic method. The color of the heatmap reflects the proportions of between-locations Markov jumps to the total jumps in the distinct time period. The directions supported by Bayesian factor (BF) are highlighted by black (>20) or red (>150) edges. Results of a full genomic data set (A), along with two data sets with reduced samples from county YL are shown (B and C). Using the same genomic data as panel (A), Bayesian inference was performed for panel (D) with geographical states assigned independently of outbreak records. The locations were determined by the sequence metadata if available, or using a uniform prior over all isolated counties/cities if the collection site was unknown.

**Supplementary figure 5.**
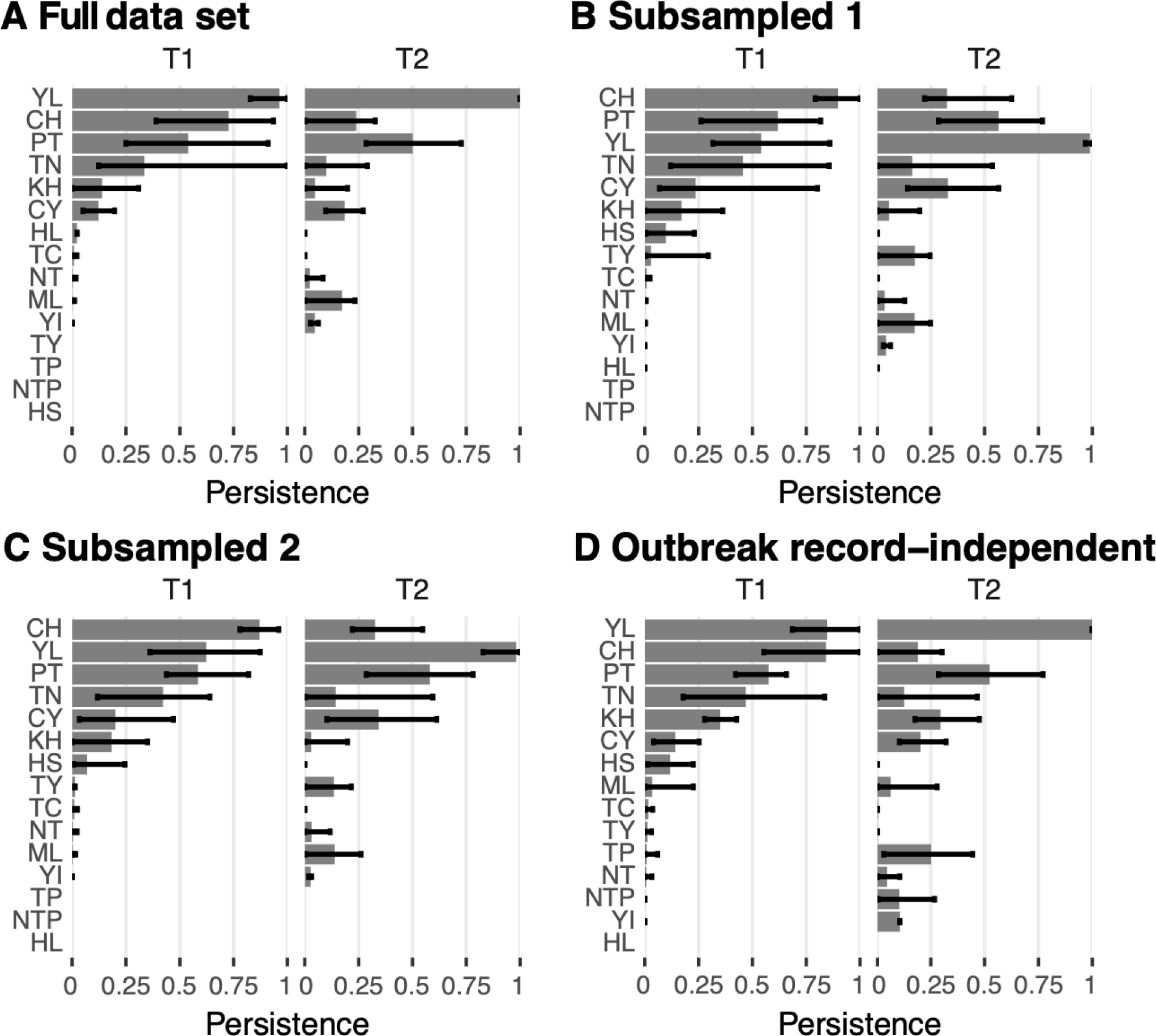
Persistence of the virus assessed by the discrete phylogeographic method. To calculate the value for each location, the branch length was unified from branches where both nodes were estimated to share the same state. The unified time interval was presented as a proportion divided by the time span of T1/T2. The mean and 95% HPD of the proportions summarized by 1000 posterior trees are indicated by the bars. The results of the four panels were obtained from the same MCMC runs as the corresponding panels in Supplementary Figure 4.

**Supplementary figure 6.**
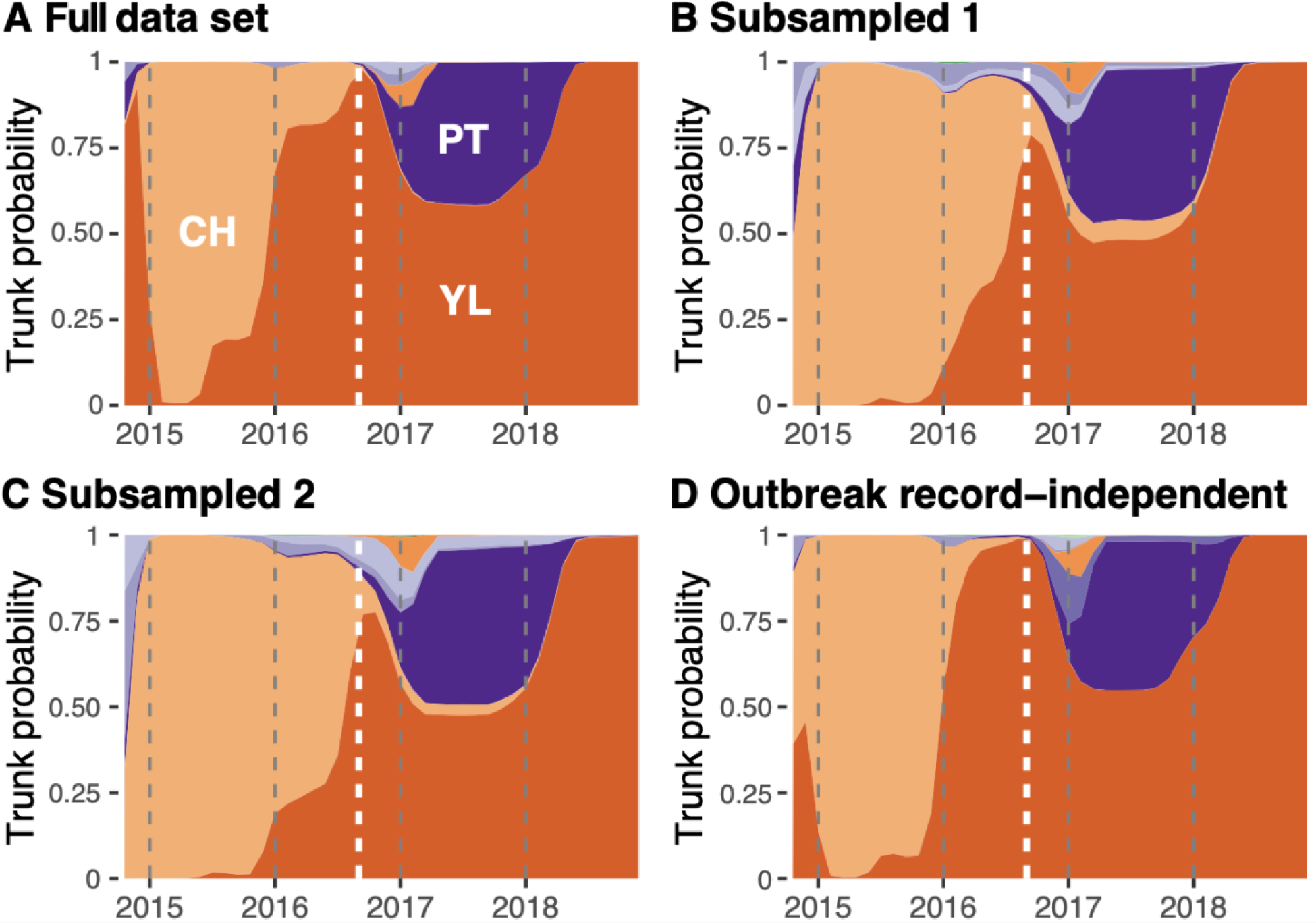
Inferred trunk locations of Taiwan clade 2.3.4.4c phylogenies through time in different genetic data sets (A-C) or with a simple geographic state assignment scheme (D). The proportions at each time point indicate the posterior support for viruses circulating in a particular county/city occupying the trunk of the tree. The areas are colored using the same scheme as Figure 2. The white dashed lines indicate the boundary of T1 and T2, September 1st, 2016.

**Supplementary figure 7.**
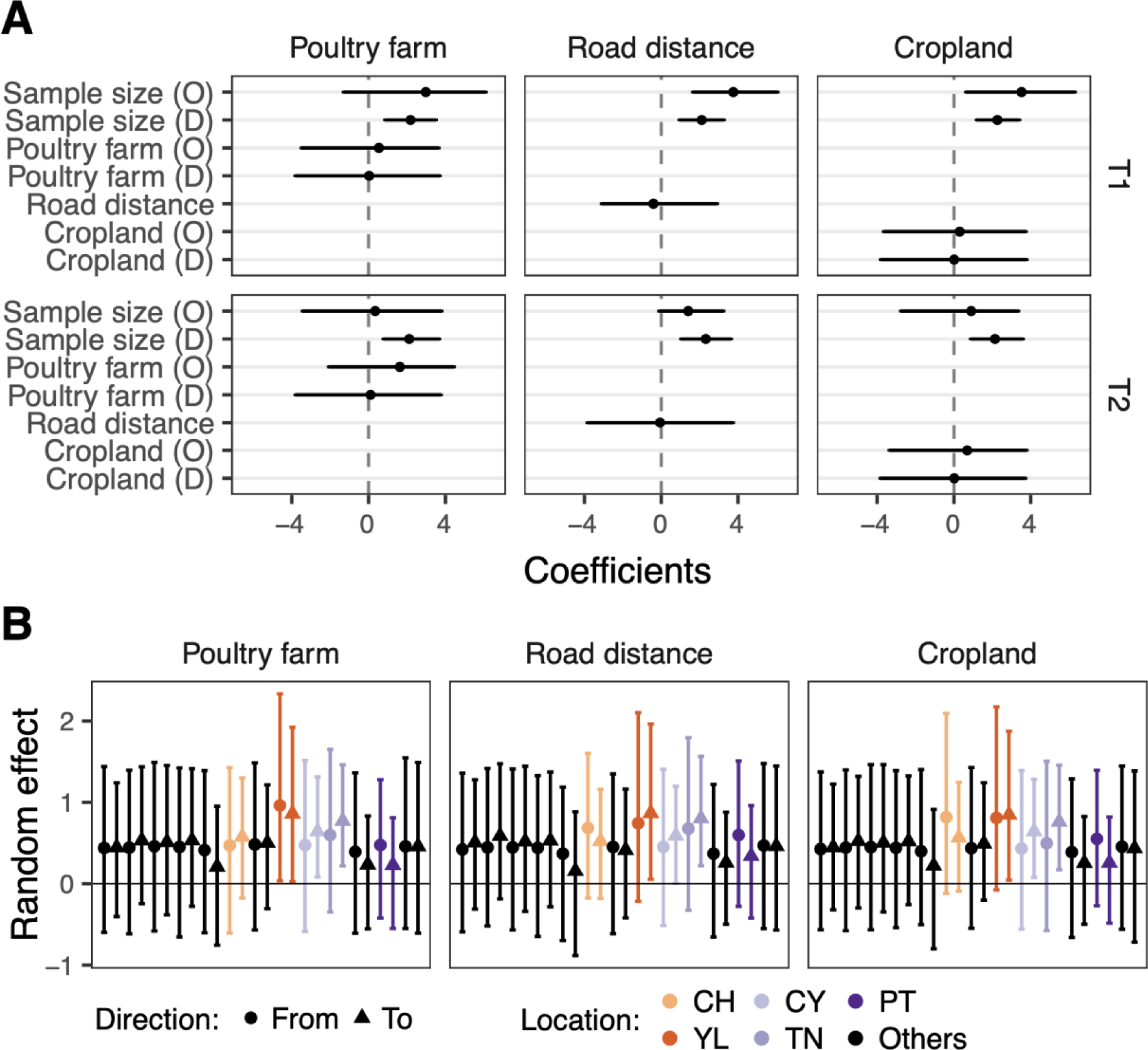
Evaluating predictors in reduced models of time-heterogeneous phylogenetic GLM. (A) The conditional effect sizes in models containing respective predictors. The predictor names are denoted by O in parentheses for origin and D for destination. (B) Location-specific random effects in the three models, with high incidence areas colored. The estimates are in log space and presented as mean with 95% HPD interval.

**Supplementary figure 8.**
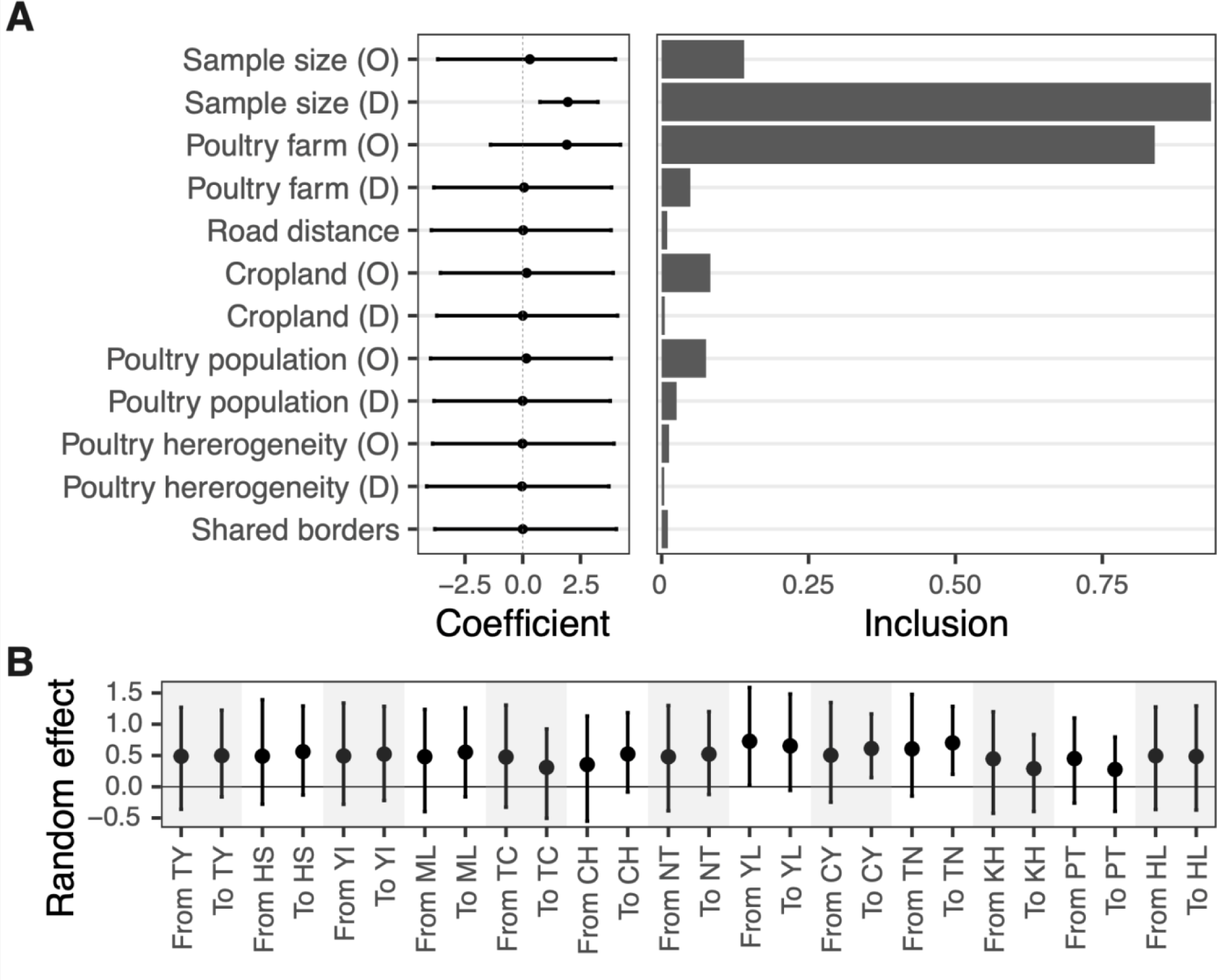
Evaluating predictors in a time-homogeneous model. (A) The conditional effect sizes and the inclusion probabilities of predictors estimated by the conventional phylogenetic GLM models. The predictor names are denoted by O in parentheses for origin and D for destination. (B) Location-specific random effects in the GLM model. The estimates are in log space and presented as mean with 95% HPD interval.

**Supplementary figure 9.**
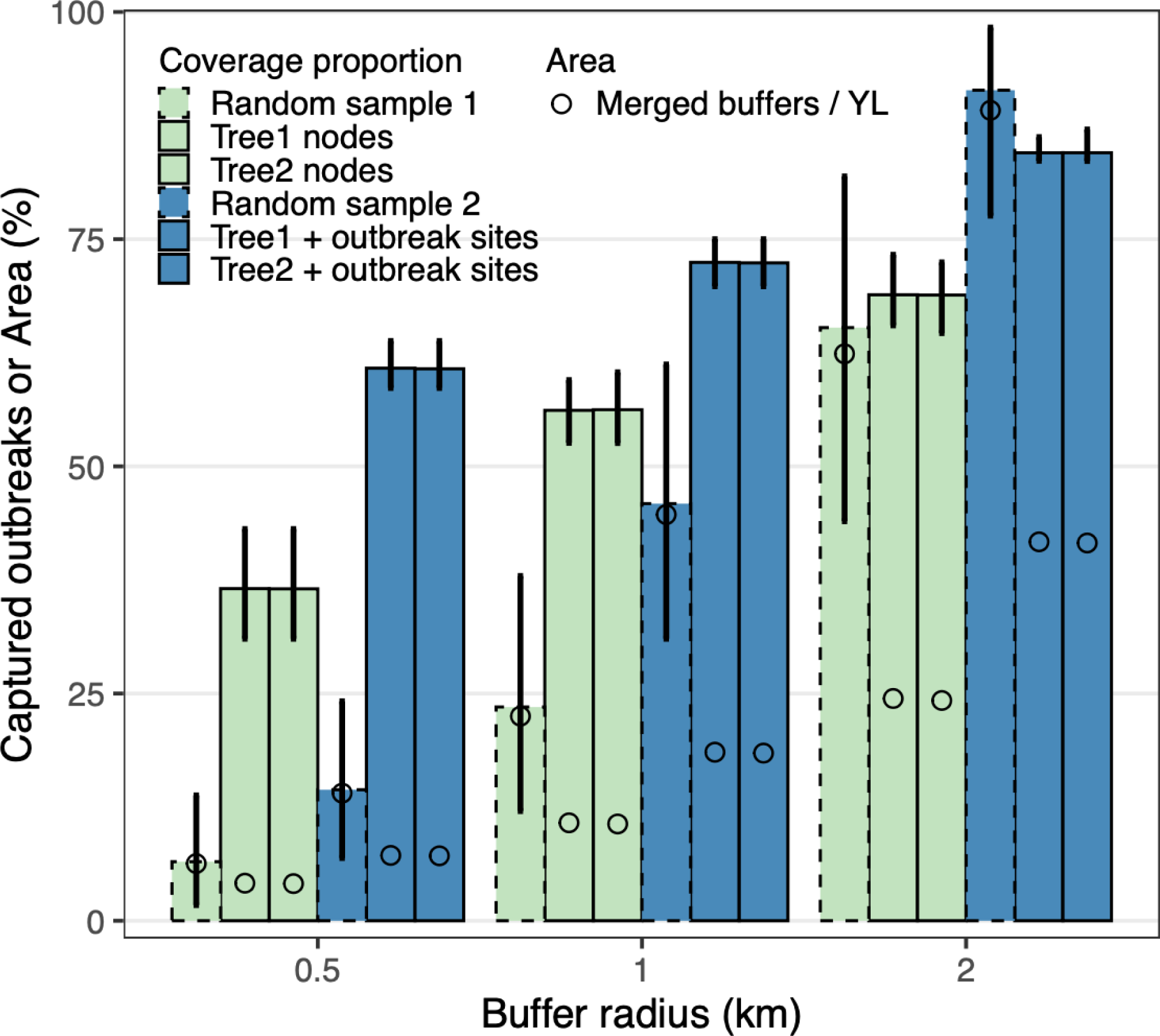
Impact of buffer radius on evaluating the re-emergence of new outbreaks in county YL. The estimated locations of tree nodes using the continuous phylogenetic method during T2, combined with or without contemporary outbreak sites, served as central points to create buffers with different radius distances on the map. The proportion of new outbreak sites covered by the buffer areas was calculated for each posterior tree. These outbreak sites were reported between 2019 and 2022, after the latest available genetic data. Null models were created by randomly distributing sites in YL with the same number of tree nodes (Random sample 1, green) or tree nodes plus outbreak sites (Random sample 2, blue). The bars show the mean values of two parallel continuous phylogeographic analyses each with 1000 posterior trees, and null models. Error bars indicate 95% credible intervals. The open circle in each bar represents the ratio of the area of merged buffers to the area of YL.

**Supplementary figure 10.**
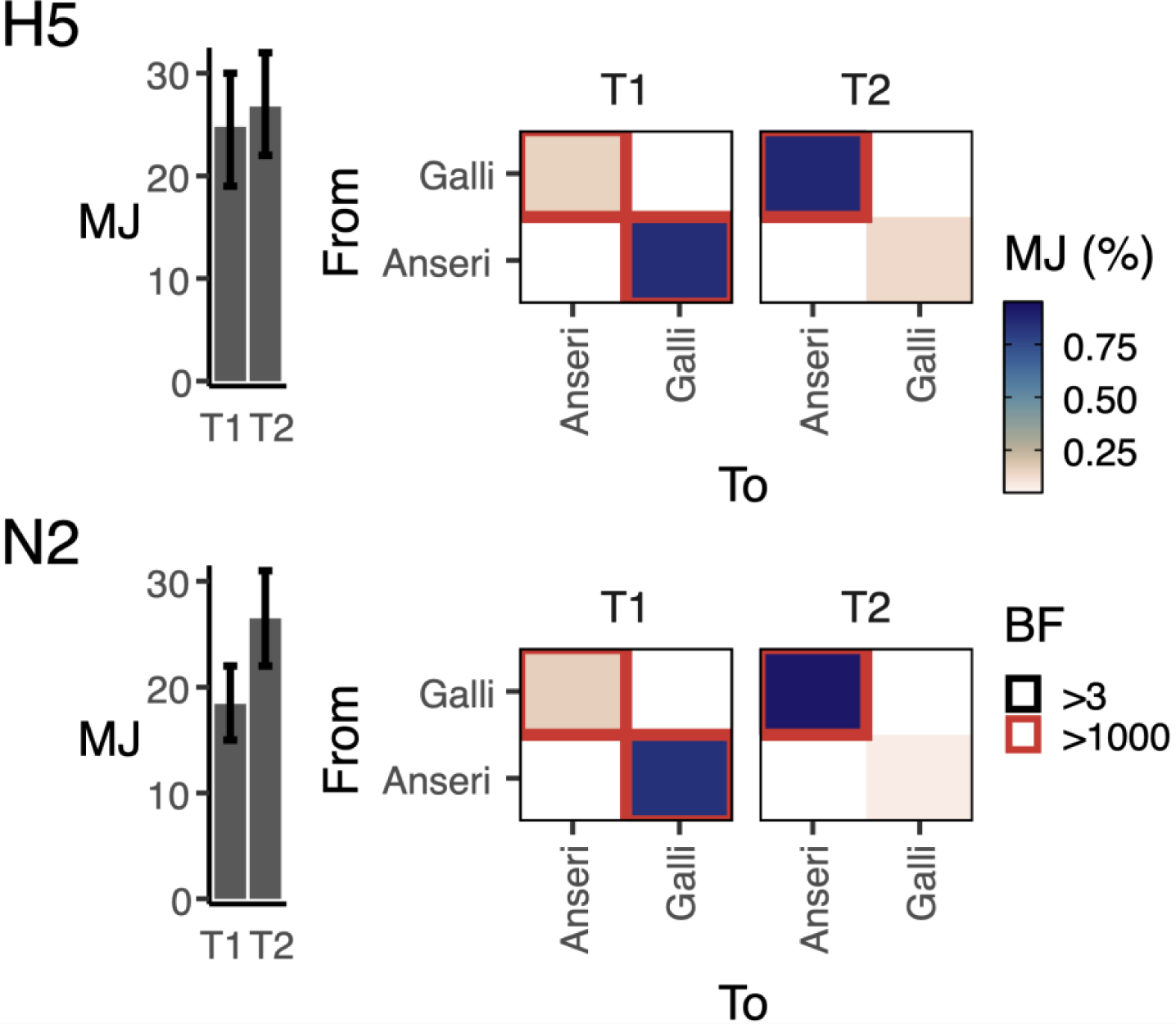
Diffusion between host groups. Markov jumps (MJ) and Bayes factors (BF) were estimated by the discrete phylogenetic method. Total MJ counts in different time spans are shown on the left. The heatmaps present the jumps as proportions to the total counts. Anseri refers to the Anseriformes genus and Galli refers to the Galliformes genus. Results based on both HA (H5) and NA (N2) are shown.

**Supplementary Table1.** Selection pressure for the surface proteins of H5 avian influenza lineages in Taiwan.

|  | H5 |  | N2 |  |
| --- | --- | --- | --- | --- |
|  | GsGd<br>Clade 2.3.4.4 | North<br>American | GsGd<br>Clade 2.3.4.4 | North<br>American |
| dN/dS ($\omega$)<br>(95% CI) | 0.204<br>(0.180-0.230) | 0.207<br>(0.174-0.245) | 0.355<br>(0.316-0.397) | 0.233<br>(0.192-0.279) |
| Pervasive positively<br>selected sites | 131 | - | 77 | - |
| Episodic positively<br>selected sites | 131, 387, 403, 545 | 17, 102, 137,<br>155, 317 | 77, 313, 416, 439 | 5, 6, 261, 307 |
| No. of fixed sites<br>among selected sites | 0 | 0 | 0 | 0 |

**Supplementary Table 2.**
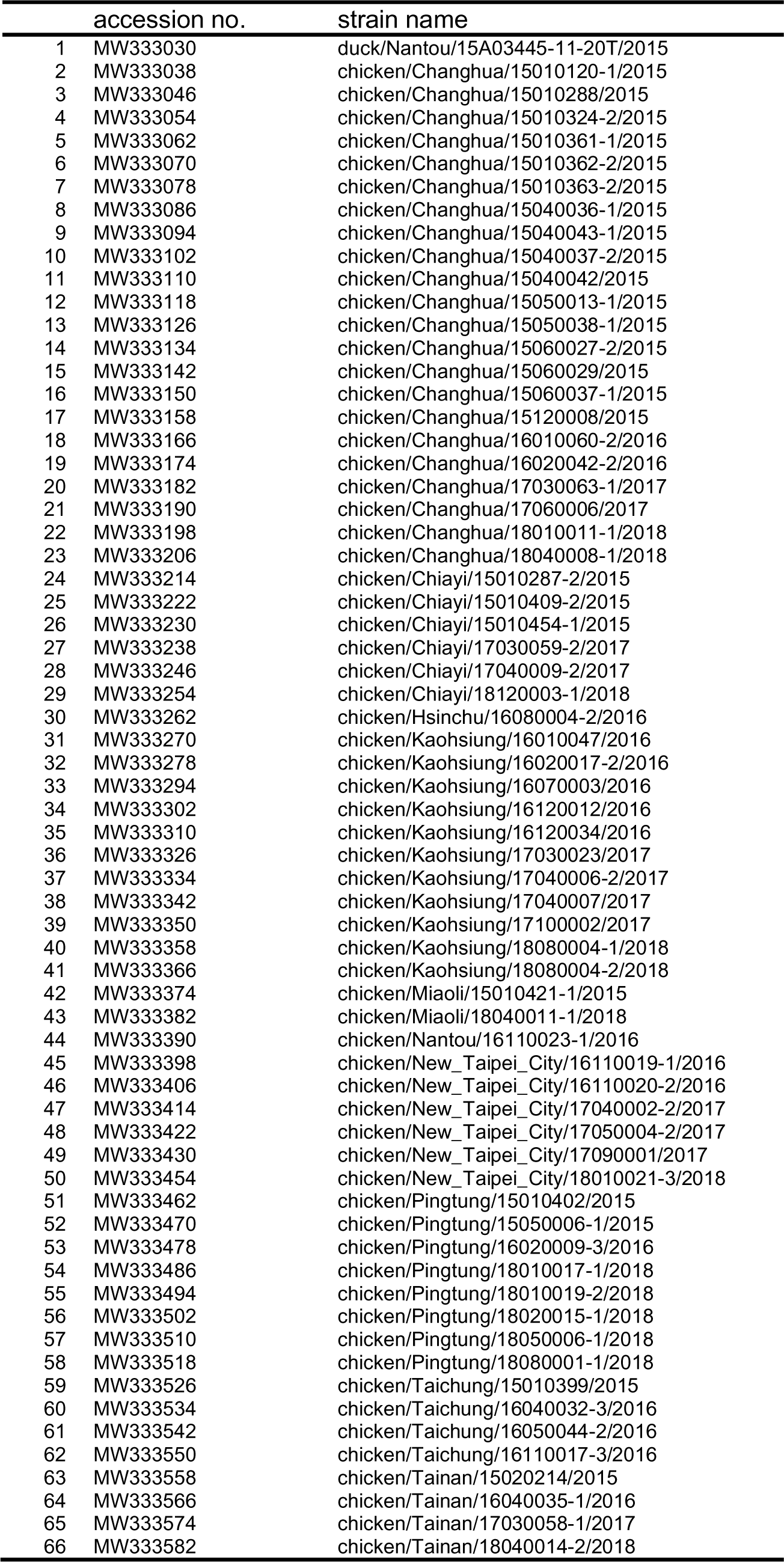
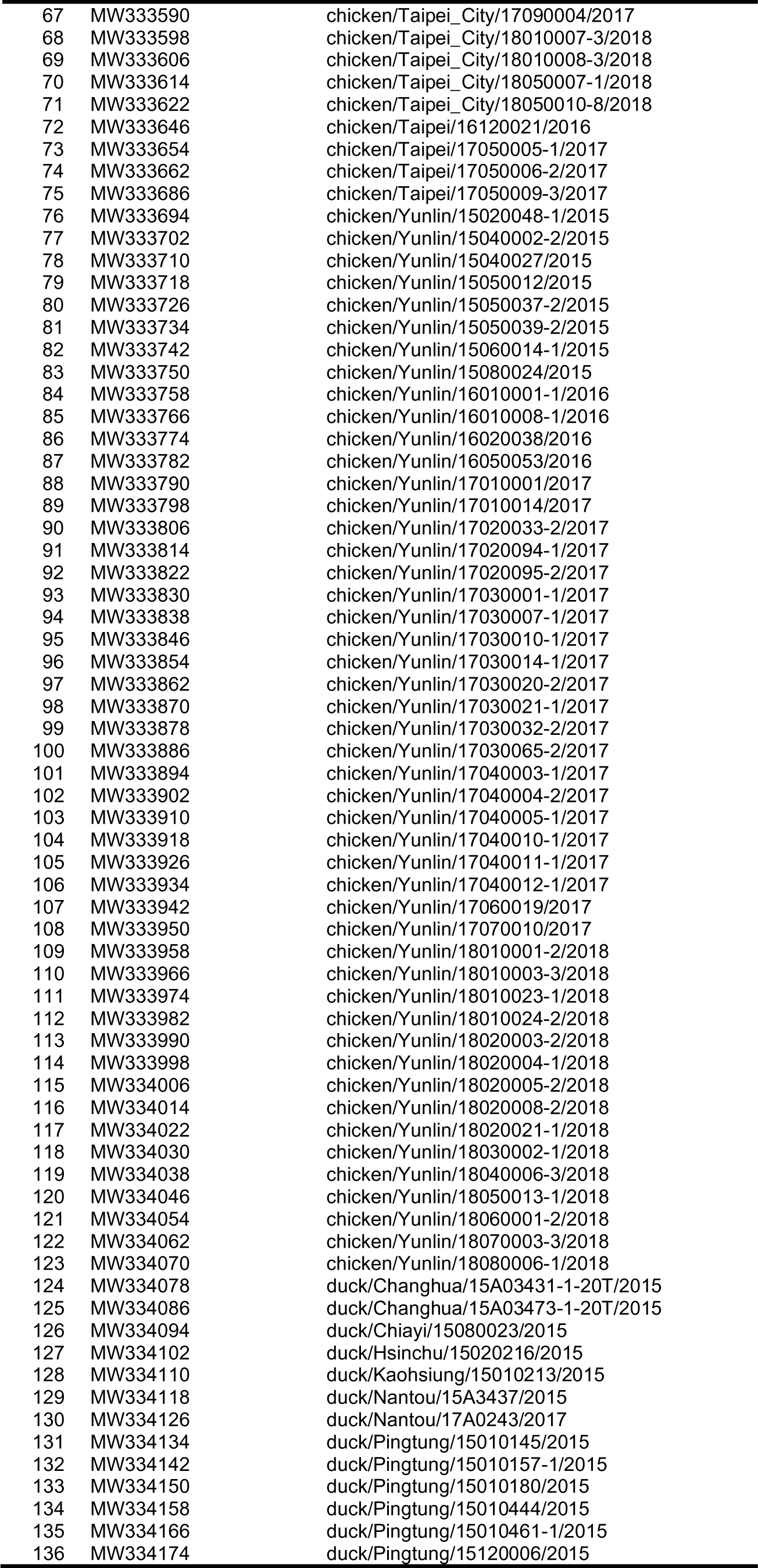
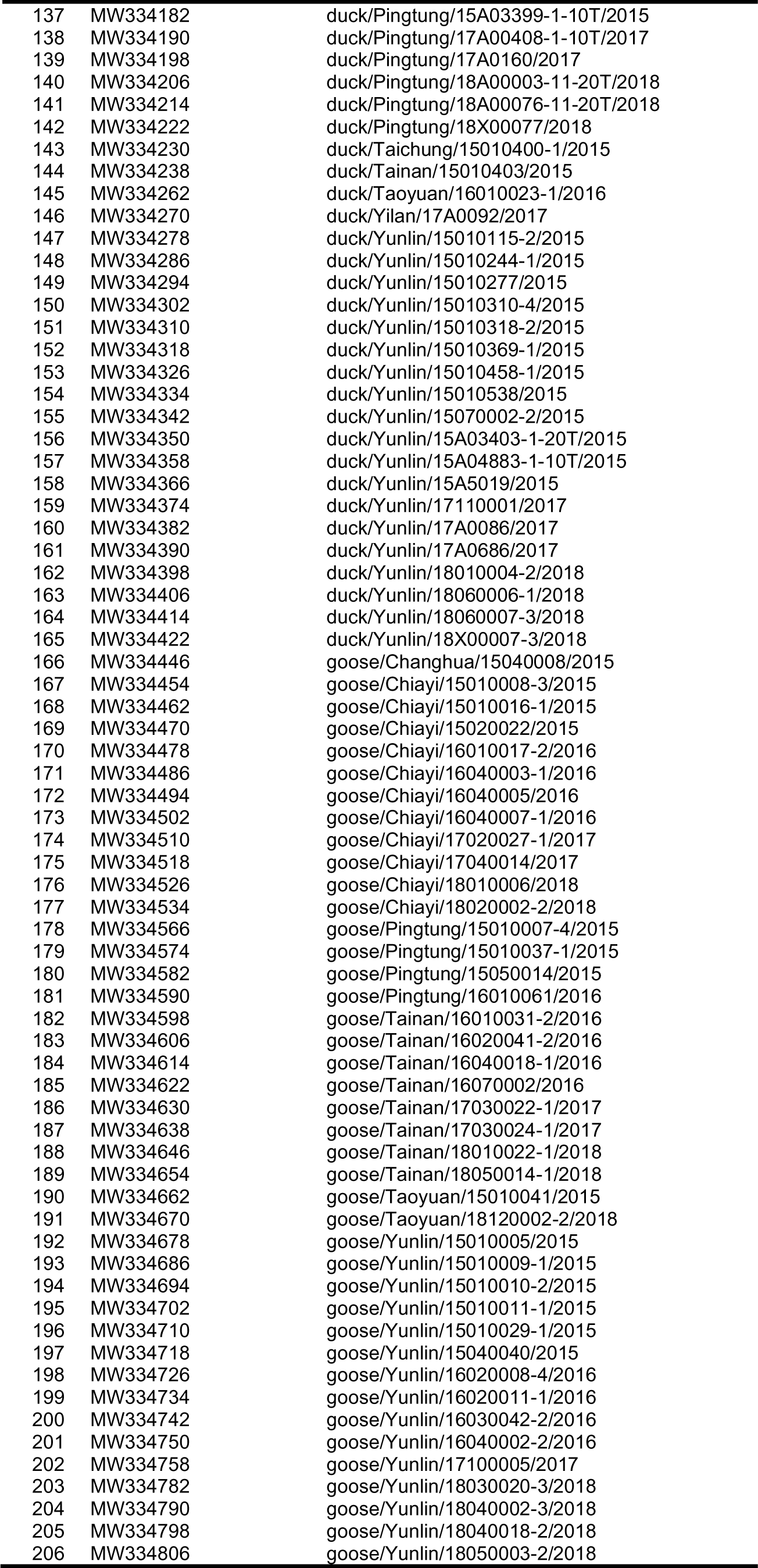
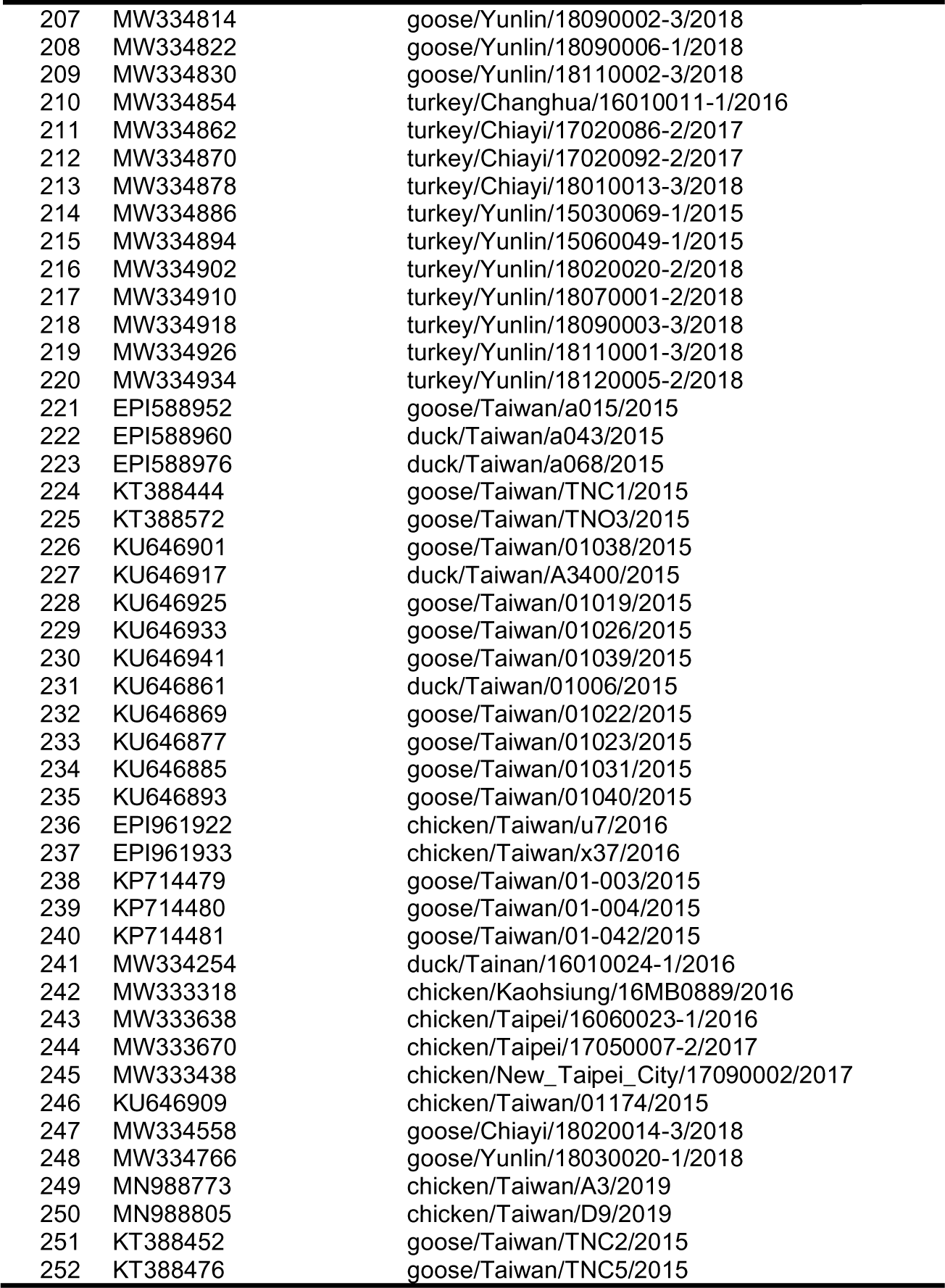
List of database accession numbers from GenBank and GISAID.

